# Influence of climatic and environmental risk factors on child diarrhea and enteropathogen infection and predictions under climate change in rural Bangladesh

**DOI:** 10.1101/2022.09.26.22280367

**Authors:** Jessica A. Grembi, Anna T. Nguyen, Marie Riviere, Gabriella Barratt Heitmann, Arusha Patil, Tejas S. Athni, Stephanie Djajadi, Ayse Ercumen, Audrie Lin, Yoshika Crider, Andrew Mertens, John M. Colford, Benjamin F. Arnold, Md Abdul Karim, Md Ohedul Islam, Rana Miah, Syeda L. Famida, Md Saheen Hossen, Palash Mutsuddi, Shahjahan Ali, Md Ziaur Rahman, Zahir Hussain, Abul K. Shoab, Rashidul Haque, Mahbubur Rahman, Leanne Unicomb, Stephen P. Luby, Adam Bennett, Jade Benjamin-Chung

**Affiliations:** Division of Infectious Diseases and Geographic Medicine, Department of Medicine, School of Medicine, Stanford University, Stanford, CA, USA; Department of Epidemiology and Population Health, School of Medicine, Stanford University, Stanford, CA, USA; Division of Epidemiology, School of Public Health, University of California, Berkeley, Berkeley, CA, USA; Department of Forestry and Environmental Resources, NC State University, Raleigh, NC, USA; King Center on Global Development, Stanford University, Stanford, California, USA; Francis I. Proctor Foundation and Department of Ophthalmology, University of California, San Francisco, San Francisco, CA, USA; Infectious Disease Division, International Centre for Diarrhoeal Disease Research, Bangladesh, Dhaka, Bangladesh; Malaria Elimination Initiative, Global Health Group, University of California San Francisco, San Francisco, CA, USA; Chan Zuckerberg Biohub, San Francisco, CA, USA

## Abstract

**Background:** Understanding pathogen-specific relationships with climate is crucial to informing interventions under climate change.

**Methods:** We matched spatiotemporal temperature, precipitation, surface water, and humidity data to data from a trial in rural Bangladesh that measured diarrhea and enteropathogen prevalence in children 0-2 years from 2012-2016. We fit generalized additive models and estimated percent changes in prevalence using projected precipitation under Shared Socio-Economic pathways describing sustainable development (SSP1), middle of the road (SSP2), and fossil fuel development (SSP5) scenarios.

**Findings:** An increase from 15°C to 30°C in weekly average temperature was associated with 5.0% higher diarrhea, 6.4% higher Norovirus, and 13.0% higher STEC prevalence. Above-median precipitation was associated with 1.27-fold (95% CI 0.99, 1.61) higher diarrhea; higher *Cryptosporidium*, tEPEC, ST-ETEC, STEC, *Shigella*, EAEC, Campylobacter, *Aeromonas*, and Adenovirus 40/41; and lower aEPEC, *Giardia*, Sapovirus, and Norovirus prevalence. Other associations were weak or null. Compared to the study period, diarrhea prevalence was similar under SSP1 (7%), 3.4% (2.7%, 4.3%) higher under SSP2, and 5.7% (4.4%, 7.0%) higher under SSP5. Prevalence of pathogens responsible for a large share of moderate-to-severe diarrhea in this setting (*Shigella, Aeromonas)* were 13-20% higher under SSP2 and SSP5.

**Interpretation:** Higher temperatures and precipitation were associated with higher prevalence of diarrhea and multiple enteropathogens; higher precipitation was associated with lower prevalence of some enteric viruses. Under likely climate change scenarios, we projected increased prevalence of diarrhea and enteropathogens responsible for clinical illness. Our findings inform pathogen-specific adaptation and mitigation strategies and priorities for vaccine development.

**Funding:** Bill & Melinda Gates Foundation, National Institute of Allergy and Infectious Diseases, National Heart, Lung, And Blood Institute, Stanford University School of Medicine, Chan Zuckerberg Biohub

**Research in Context:** *Evidence before this study:* We searched Google Scholar and Scopus for studies published from January 1, 2000 to present using the following three queries: 1) child; and diarrhea OR “enteric infection”; and meteorological OR environmental OR “surface water” OR “standing water”; and risk AND/OR factors; 2) Climate AND change AND project* AND diarrhea OR diarrhoea; 3) climate AND change AND project* OR model AND enter* AND infect* AND E. coli. Studies generally focused on individual risk factors for diarrhea transmission or enteric infection, with an emphasis on temperature and precipitation. Studies found that higher temperatures were associated with higher incidence of bacterial diarrhea and lower incidence of viral diarrhea; few studies have investigated associations between temperature and parasitic diarrhea. Heavy rainfall, particularly after dry periods, was associated with higher diarrhea prevalence, though heavy rainfall during rainy seasons was found to protect against diarrhea incidence. Similarly, flooding of surface water and shallow wells was also associated with higher diarrhea prevalence. Very few studies investigated associations between diarrhea or enteropathogen carriage and surface water presence, or humidity. A recent individual participant meta-analysis of studies in 19 low- and middle-income countries found that higher precipitation was associated with a small decrease in enterotoxigenic *E. coli* (ETEC) and *Campylobacter* spp. prevalence and no difference in *Shigella, Cryptosporidium*, or *Giardia*, or enteric virus prevalence. Weekly average temperature increases of 10-40° C within the study period were associated with higher risk of *Campylobacter*, ETEC, *Shigella, Cryptosporidium, Giardia*, and adenovirus, and lower risk of sapovirus and rotavirus, and generally, associations were stronger. Higher humidity was associated with higher risk of enteric bacterial infections and lower risk of enteric virus infection. A small number of studies have projected diarrhea under climate change in low- and middle-income countries. Studies have estimated a 15-20% increase in global diarrhea risk in 2040-2069 relative to 1961-1990, up to 21% increase in diarrhea incidence in northern India from 2013 to the 2040s, 3-10% increase in diarrhea cases in the Gaza Strip associated with 1.5° to 2°C increases in temperature, and an 8% increase in diarrhea burden by 2050 in Nepal. One study estimated an additional 1,625,073 ETEC diarrhea cases in Bangladesh from 2046-2065 due to climate change. No studies estimated changes in the presence of multiple enteropathogens under possible climate change scenarios.

*Added value of this study:* This study matched remote sensing data on precipitation, temperature, surface water, and humidity to data on diarrhea and enteropathogen carriage in children under 2 years from a trial in rural Bangladesh. We fit flexible models to investigate potentially non-linear relationships between each climate and environmental risk factor and each health outcome. In addition, we predicted the prevalence of each outcome under three possible climate change scenarios. Our predictions leveraged the spatiotemporal distributions of both climate precipitation projections and diarrhea and enteropathogen carriage in the trial. To our knowledge, this is the first study to predict prevalence under climate change scenarios for both diarrhea and multiple enteropathogens in a rural, low-resource south Asian setting. Our study sheds light on how climate change may impact diarrhea and enteropathogen carriage in Bangladesh, which is highly vulnerable to climate change due to its low altitude and seasonal flooding.

*Implications of all the available evidence:* We found that both higher temperatures and precipitation were associated with higher diarrhea prevalence, consistent with prior studies. Compared to prior studies, which have primarily found associations between diarrhea and heavy rainfall following a dry period, we found that moderate levels of weekly precipitation were associated with higher diarrhea prevalence. Relationships between climatic and environmental variables and enteropathogen carriage varied by taxa, as other studies have found, but for some pathogens and risk factors, we observed associations in different directions than prior studies. Under middle of the road and fossil fuel-based development climate change scenarios, we projected increased diarrhea prevalence and taxa-specific changes in enteropathogen prevalence. Our findings suggest that climate change may exacerbate diarrhea burden in rural Bangladesh in the absence of sustainable development and inform prioritization of pathogen-specific mitigation and/or adaptation interventions (e.g., vaccines) for young children in rural, South Asian settings under climate change.

## Introduction

Diarrhea was the third leading cause of disability adjusted life years (DALYs) among children under 10 years in 2019.^1^ Among young children, diarrhea can result in long-term impacts on growth^2^ and cognition.^3^ Where diarrhea burden is high, asymptomatic enteropathogen carriage is common^4^ and is linked to child growth failure^4^ and impaired child cognitive development.^5^ Climatic and environmental factors may influence diarrhea and enteropathogen carriage in diverse ways due to pathogens’ differing environmental survival and transport.^6^ Prior studies have found increased diarrhea after heavy rainfall and floods^7,8^ and that parasitic and bacterial diarrhea are more common in the rainy season.^9^ Increased temperatures have been associated with higher incidence of bacterial diarrhea but lower incidence of viral diarrhea.^9^ In low- and middle-income countries (LMICs), higher temperatures have been associated with a decrease in rotavirus prevalence and small increase in adenovirus prevalence; higher humidity and temperature have been associated with higher enteric bacteria prevalence and lower virus prevalence; and associations with precipitation varied by pathogen.^10^

A small number of studies have estimated the burden of diarrhea and enteropathogen infection under climate change.^11–16^ The World Health Organization estimated an additional 32,955 diarrhea deaths in children under 15 years in 2050 due to climate change, with the greatest increases in South Asia and Eastern Africa.^17^ Another study projected that diarrhea risk would increase by 15-20% in the period 2040-2069 relative to risk levels in 1961-1990.^11^ Since the etiology of diarrhea varies by age and geography,^18^ understanding pathogen-specific relationships with climatic variables at present and with climate change can inform adaptation and mitigation strategies.^8,11,17^ Enteropathogens whose burden is most likely to increase under climate change could be prioritized for vaccine development. However, no studies have projected the impact of climate change on the prevalence of multiple enteropathogens.

Our objectives were to identify climatic and environmental risk factors associated with diarrhea and enteropathogen prevalence in young children in rural Bangladesh and to predict changes in diarrhea and enteropathogen prevalence under potential climate change scenarios.

## Methods

### Study design

We analyzed data that was collected in a cluster-randomized trial of water, sanitation, handwashing, and nutrition interventions in rural Gazipur, Mymensingh, Tangail, and Kishoreganj districts of Bangladesh.^19^ From 2012-2013, the trial enrolled a total of 720 village clusters spanning a geographic area of 12,500 km^2^. The trial enrolled pregnant women and measured outcomes in their children at enrollment and approximately 1 and 2 years later. We matched climatic and environmental risk factors from remote sensing datasets to trial data by date of outcome measurements and geocoordinates of study compounds (groups of households in which patrilineal families share a common courtyard). Our pre-analysis plan is available at https://osf.io/f9cza/, and we note deviations from it in Appendix 1.

### Outcomes

Field workers measured caregiver-reported diarrhea (three or more loose or watery stools in a 24-hour period or a single stool with blood) in children 6 months - 5.5 years old (N=4,478 children). Diarrhea prevalence was measured in three rounds (2012-3, 2013-4, 2014-5). Field workers collected stool samples from a subsample of children aged 14 months in 2014 (N=1,645 children) (details in Appendix 2) and detected a panel of 34 enteric viruses, bacteria and parasites in stool using qPCR (N=1,408 children’s samples) (details in Appendix 3).

We measured the following pre-specified primary outcomes: prevalence of 1) caregiver-reported diarrhea in the past 7 days, 2) any enteric virus (adenovirus 40/41, astrovirus, norovirus GI/GII, rotavirus, sapovirus), and 3) any parasite (*Cryptosporidium* spp, *Enterocytozoon bieneusi*, or *Giardia* spp). We did not include prevalence of any bacteria because at least one bacterium was detected in over 95% of samples. Pathogen-specific prevalence for pathogens detected in >10% of samples were secondary outcomes. To detect potential misclassification of diarrhea, we included caregiver-reported child bruising in the past 7 days as a negative control outcome.

### Climatic and environmental variables

Here we briefly summarize each variable; Appendix 4 includes additional details.

#### Temperature

Using 1 km resolution daily temperature data from the National Aeronautics and Space Administration (NASA),^20^ we calculated the average, minimum and maximum temperature values for 7 or 30 days prior to observation with 1-3 week lags for diarrhea and 0-3 week lags for pathogen outcomes.

#### Precipitation

We obtained 0.5 degree resolution precipitation data from the National Oceanic and Atmospheric Administration.^21^ We created an indicator for heavy rainfall (any day in the prior week with total daily precipitation > 80^th^ percentile on rainy days during the study period) and an indicator for whether the weekly sum of precipitation was above or below the median. We included 1-3-week lags for diarrhea and 0-3-week lags for pathogen outcomes.

#### Surface water

We obtained 30 m resolution surface water data for the period 1984-2020 from the Global Surface Water Explorer.^22^ Variables included: 1) seasonal surface water consistently present in a season, 2) ephemeral surface water present intermittently, and 3) any surface water (ephemeral, seasonal, or permanent). We calculated tertiles of distance from each household to the nearest surface water and created an indicator for whether the proportion of pixels with surface water within 250m, 500m, or 750m of each household was above or below the median.

#### Humidity

We obtained mean monthly vapor pressure deficit data with 4 km resolution from Terraclimate^23^ and measured associations with a continuous measure in kilopascals (kPA).

### Statistical analysis

We restricted diarrhea analyses to the control arms of the original trial (N=7,320 measurements) because a companion analysis found that intervention effects on diarrhea were modified by climate and environment.^24^ For pathogen carriage outcomes, we pooled across all study arms in which specimens were collected (N=1,408 measurements) and included study arm as a covariate.

We used generalized additive mixed models to estimate the relationship between continuous exposures (modeled with cubic splines) and outcomes. We adjusted for covariates that were associated with each outcome in our dataset using a likelihood ratio test (p-value < 0.1) (details in Appendix 5). Because pair-wise correlations between climatic exposures were not strong (Figure A1), we did not adjust individual exposure models for other climatic exposures. To assess potential spatial autocorrelation, we used Moran’s I coefficient and included a bidimensional thin plate spline function of household latitude and longitude for models in which we detected spatial autocorrelation. We included random intercepts for study compound for diarrhea models to account for multiple measurements per child and per compound and random intercepts for study cluster for pathogen outcomes to account for village-level clustering. To estimate simultaneous confidence intervals, we resampled from the variance-covariance matrix under a multivariate normal distribution.^25^

We predicted the percentage point change in prevalence under projected precipitation in 2050 using three climate change scenarios: sustainable development reduces CO_2_ emissions to net zero by 2050 (Shared Socioeconomic Pathways 1 (SSP1)); current socioeconomic trends remain similar and progress towards sustainability is slow, and CO_2_ emissions remain the same in 2050 (SSP2); and fossil fuel-based development dominates and CO_2_ emissions double by 2050 (SSP5).^26^ We matched precipitation projections for 2050 from the Coupled Model Intercomparison Project Phase 6 data from the NASA Center for Climate Simulation to study household location and measurement week. We calculated mean weekly precipitation using 0-3 week lags (see details in Appendix 6). Because the generalized additive model spline fits were approximately linear, for computational efficiency, we fit generalized linear models with a binomial link function between outcomes and the weekly sum of precipitation then used parametric g-computation to calculate percent changes in predicted prevalence under each SSP scenario vs. the study period. We used a bootstrap with 1,000 replicates, resampling by study clusters for pathogen outcomes and by study compounds for the diarrhea analysis. Our primary analysis used precipitation lag periods that best aligned with the incubation period for each outcome (Appendix 7) and repeated analyses with alternative lag periods. We assumed that the association between precipitation and each outcome will remain constant over time.

We assessed effect modification for each outcome by child age (<1.5 vs. 1.5 years) for diarrhea outcomes. For precipitation analyses, we assessed effect modification by temperature.

### Role of the funding source

The funders of the study had no role in study design, data analysis, interpretation, writing of the manuscript, or the decision to submit the manuscript for publication.

## Results

### Study participant characteristics

The diarrhea cohort included 4,478 children (mean age = 23 months; SD = 12) measured in three rounds from 2012-2016, and the pathogen cohort included 1,408 children (mean age = 14 months; SD = 2) measured primarily in 2014 (Table 1). Other demographic characteristics of each cohort have previously been reported.^19,27^

**Table 1.** Study participant characteristics.

|  | **Diarrhea cohort** | **Pathogens cohort** |
| --- | --- | --- |
| Children | 4,478 | 1,408 |
| Observations | 7,320 | 1,408 |
| **Year of measurement** |  |  |
| 2012 | 491 (6.7%) | 0 (0.0%) |
| 2013 | 2,680 (36.6%) | 19 (1.3%) |
| 2014 | 1,868 (25.5%) | 1,389 (98.7%) |
| 2015 | 2,170 (29.6%) | 0 (0.0%) |
| 2016 | 111 (1.5%) | 0 (0.0%) |
| Mean age, months (SD) | 22.6 (11.8) | 14.0 (2.0) |
| Antibiotics consumed in the past week | --<sup>a</sup> | 233 (16.5%) |
| **Season** |  |  |
| Rainy | 3,408 (46.6%) | 620 (44.0%) |
| Dry | 3,912 (53.4%) | 788 (56.0%) |
| **Intervention arm in original trial** |  |  |
| Control | 7,320 (100.0%) | 328 (23.3%) |
| Water | 0 (0.0%) | 0 (0.0%) |
| Sanitation | 0 (0.0%) | 0 (0.0%) |
| Handwashing | 0 (0.0%) | 0 (0.0%) |
| WSH | 0 (0.0%) | 368 (26.1%) |
| Nutrition | 0 (0.0%) | 352 (25.0%) |
| Nutrition + WSH | 0 (0.0%) | 360 (25.6%) |

<sup>a</sup> Not measured

### Temporal trends

The study period spanned three rainy seasons from 2013-2015. During the rainy season, average weekly temperature was higher (Figure 1). Distance to surface water was slightly lower during the peak of each rainy season. Diarrhea measurements were not collected in some weeks of the 2013-14 rainy seasons.

**Figure 1.**
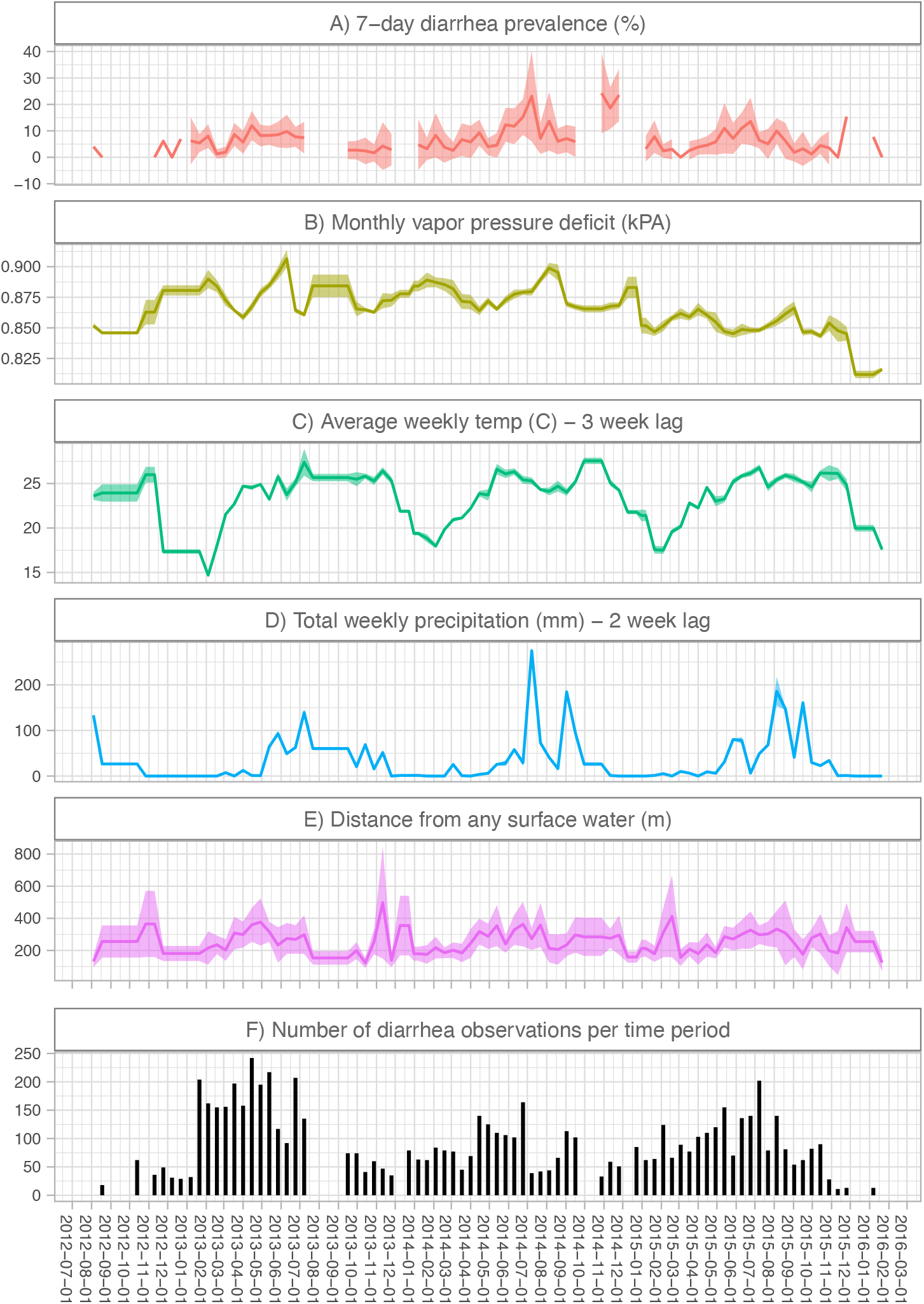
Time trends in risk factors, diarrhea prevalence, and number of observations. Shaded bands indicate 95% confidence intervals using robust sandwich standard errors to account for clustering within study clusters. Estimates excludes biweekly periods when the number of observations was <10. Panel A) includes diarrhea measurements from the control arms of the original trial. Breaks in the line indicate periods when the study did not collect data on diarrhea status. In panels B) to E), we display the risk factor values that corresponded to the diarrhea measurements in our analysis. For periods when there was no diarrhea data collection, we display the risk factor values for the next biweekly period with at least diarrhea measurements. Panel G) displays the number of diarrhea observations in each biweekly period.

### Temperature

During the study period, the weekly average temperature was 17-32°C, the weekly minimum temperature was 6-25°C, and the weekly maximum temperature was 22-40°C. Diarrhea prevalence was 2.8% (95% CI 1.0%, 7.6%) at weekly average temperature of 15°C and 7.3% (95% CI 3.2%, 15.8%) at 30° C using a 3-week lag; there was no association with shorter lags (Figure 2a). Higher average temperatures were also associated with higher norovirus prevalence (12.0% (95% CI 6.6%, 20.8%) at 17°C; 18.3% (95% CI 11.9%, 27.0%) at 30°C; 1-week lag), and higher STEC prevalence (5.2% (95% CI 1.9%, 13.8%) at 18°C; 17.8% (95% CI 9.6%, 30.5%) at 30°C; 1-week lag) (Figure 2b). There was no association with prevalence of other bacteria (Figure 2c) or parasites (Figure 2d). Overall, results for pathogen prevalence were similar using alternative lag periods (Figure A2). Diarrhea and pathogen prevalence were not meaningfully associated with weekly minimum and maximum temperature (Figure A3-5).

**Figure 2.**
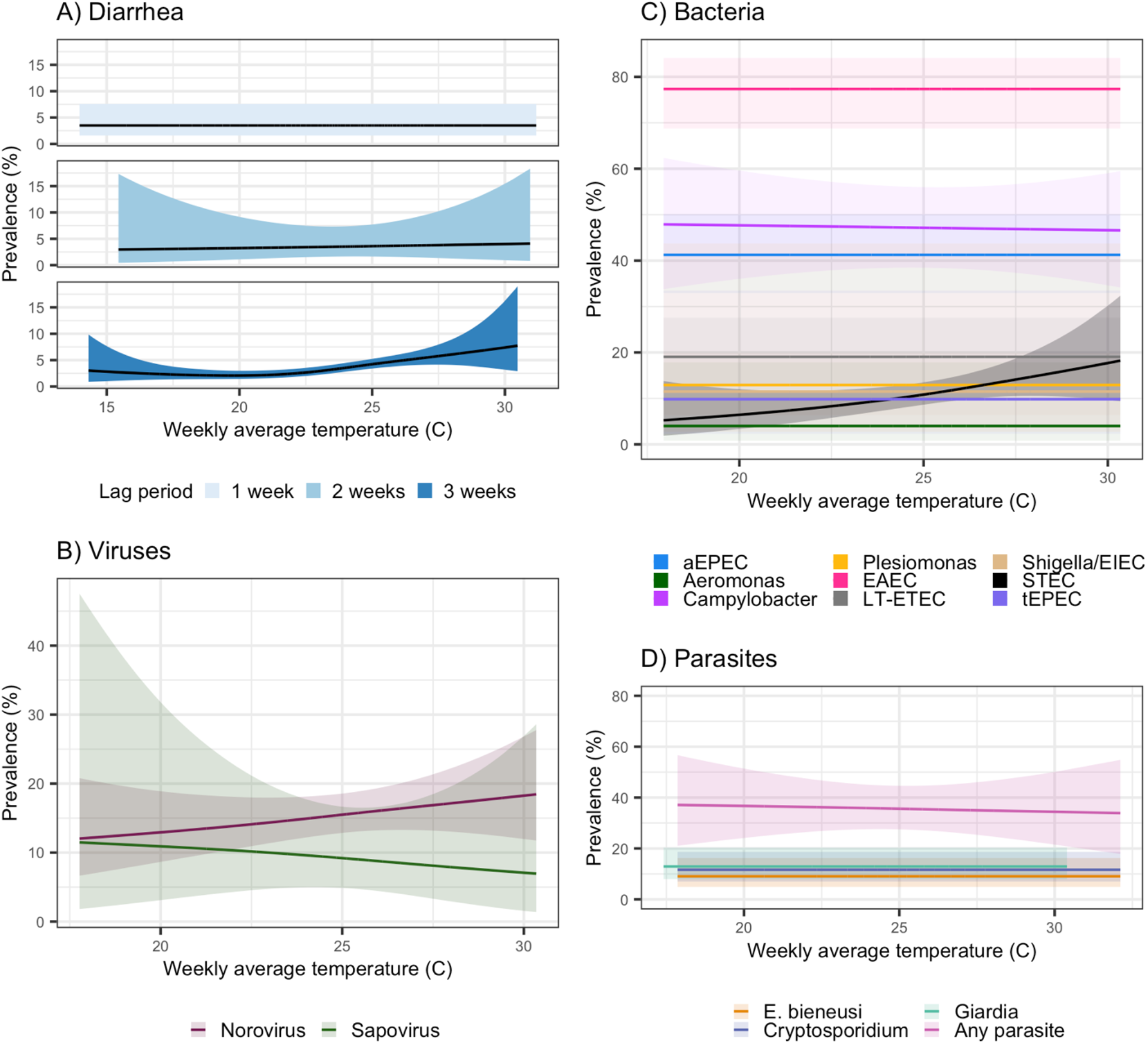
Diarrhea and enteropathogen prevalence by weekly average temperature. All panels present unadjusted models; shaded bands indicate simultaneous 95% confidence intervals accounting for clustering. Panel A) includes diarrhea measurements in children aged 6 months - 5.5 years in the control arms in the original trial. Panels B-D) include measurements in children approximately 14 months of age in the control, combined water + sanitation + handwashing (WASH), nutrition, and combined nutrition + WASH arms of the original trial. We present results for the lag periods that best aligned with the incubation period for each outcome (Appendix 7); results with alternative lag periods are in Figure A2. Results not shown for any virus, adenovirus 40/41, or ST-ETEC due to very low precision of model estimates.

### Precipitation

Mean total weekly precipitation was 52mm (range: 0-284mm) annually, 13mm (range: 0-171mm) in the dry season and 91mm (2-307mm) in the wet season. Above-median weekly precipitation was associated with higher diarrhea prevalence (PR = 1.27; 95% CI 0.99, 1.61; adjusted PR (aPR) = 1.35, 95% CI 1.10, 1.67) using a 2-week lag, but not using 1- or 3-week lags (Figure 3a). For parasites, above-median weekly precipitation was associated with higher *Cryptosporidium* prevalence (e.g., PR = 2.21; 95% CI 1.49, 3.26 for a 3-week lag), but lower *Giardia* prevalence (e.g., PR = 0.63; 95% CI 0.45, 0.89 for a 2-week lag) (Figure 3b). For viruses, above-median weekly precipitation was associated with higher prevalence of adenovirus 40/41, but lower prevalence of sapovirus and norovirus (Figure 3c). For bacteria, above-median weekly precipitation was associated with higher prevalence of typical enteropathogenic *Escherichia coli* (tEPEC), enterotoxigenic *Escherichia coli* with heat-stable toxin (ST-ETEC), Shiga toxin-producing *Escherichia coli* (STEC), *Shigella*/enteroinvasive *Escherichia coli (Shigella*/EIEC), *Plesiomonas*, enteroaggregative *Escherichia coli* (EAEC), *Campylobacter*, and *Aeromonas* and lower prevalence of atypical enteropathogenic *Escherichia coli* (aEPEC) (Figure 3d). Results varied by lag period. Heavy rainfall was associated with lower prevalence of diarrhea (PR = 0.72, 95% CI 0.54, 0.96; 2-week lag) and *E. bieneusi*, norovirus, and sapovirus, and higher prevalence of *Aeromonas, Campylobacter, Shigella*/EIEC, EAEC, and ST-ETEC (Figure A6).

**Figure 3.**
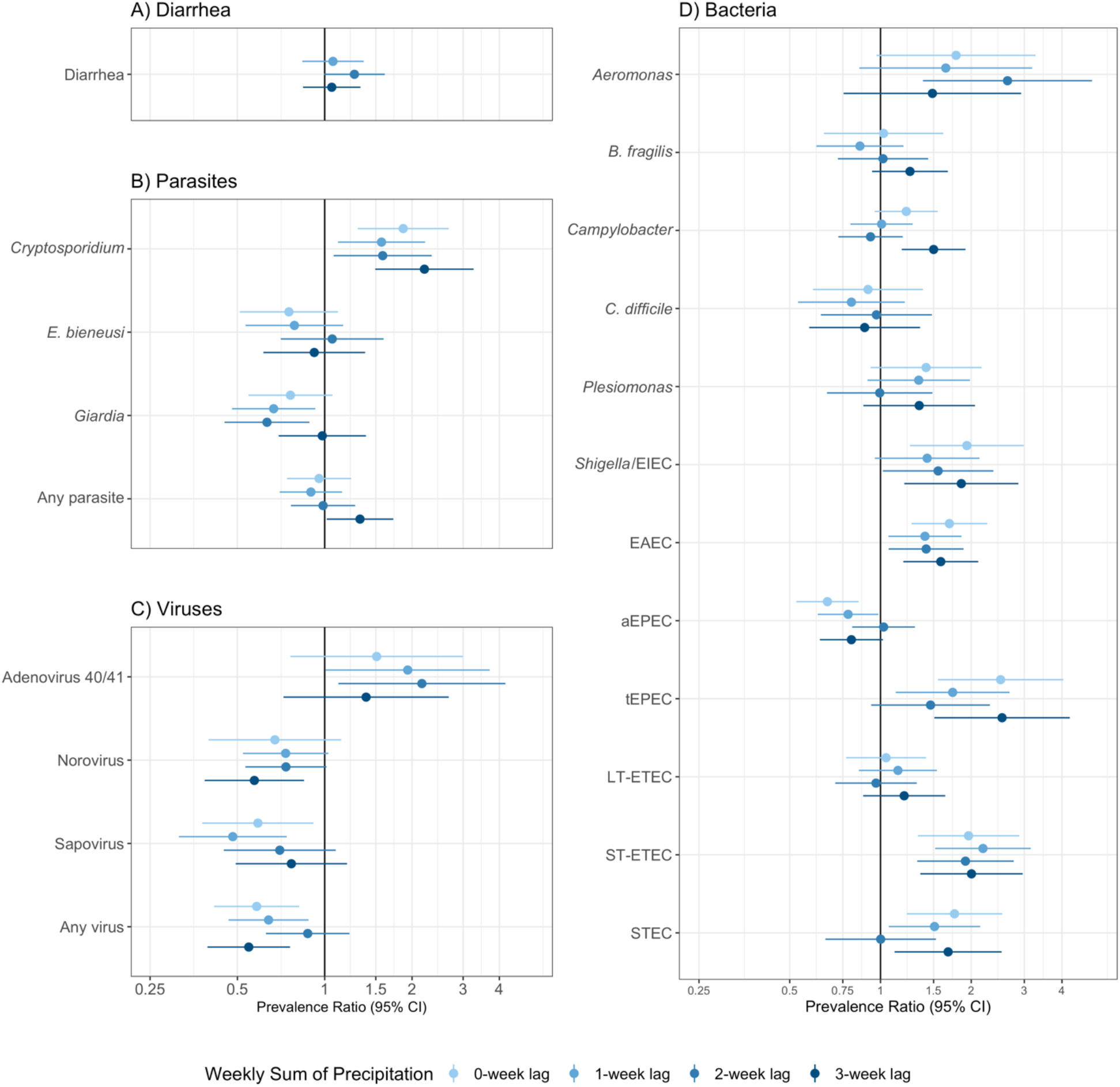
Diarrhea and enteropathogen prevalence by average weekly precipitation. All panels present unadjusted models including an indicator for above median average weekly precipitation as the independent variable; adjusted models produced similar results. Error bars present 95% confidence intervals adjusted for clustering. Panel A) includes diarrhea measurements in children aged 6 months - 5.5 years in the control arms in the original trial. Panels B-D) include measurements in children approximately 14 months of age in the control, combined water + sanitation + handwashing (WASH), nutrition, and combined nutrition + WASH arms of the original trial.

### Surface water

The mean distance from study households to the closest surface water was 287m (range: 10-1818m) for any surface water, 451m (range: 11-2083m) for ephemeral surface water, and 405m (range: 11-1902m) for seasonal surface water. Compared to households in the highest tertile of distance to any surface water (> 316m), *Aeromonas* prevalence was lower for those in the middle tertile (165-316m) (PR=0.53; 95% CI 0.29, 0.96) and in the lowest tertile (<165m) (PR=0.43; 95% CI 0.23, 0.81) (Figure A7). *Aeromonas* results were similar for ephemeral and seasonal surface water. Distance to surface water was not associated with other outcomes.

Around each study household, the median proportion of land that contained any surface water was 0.4% (range: 0-76%) within 250m, 3% (range: 0-86%) within 500m, and 5% (range: 0-84%) within 750m. Most outcomes were not associated with the proportion of land that contained surface water. For certain radii and surface water types, an above-median proportion of surface water was associated with lower diarrhea, *Aeromonas*, and adenovirus 40/41 prevalence and higher prevalence of tEPEC and *Shigella*/EIEC; however, most confidence intervals were close to or spanned the null (Figure A8).

### Vapor pressure deficit

Vapor pressure deficit (VPD) ranged from 0.57 to 1.51 kPa during the study period. Higher values of VPD, corresponding to lower humidity, were associated with small decreases in *Cryptosporidium* and EAEC (Figure A9). While there were some associations with other outcomes, precision was low.

### Projected prevalence under climate change

During the study period, total annual rainfall was 2,032 mm during the study period and 1,754 mm under SSP1, 3,329 mm under SSP2, and 3,782 mm under SSP5 in 2050 (Table A1). There were 185 rainy days during the study vs. 154 under SSP1, but total precipitation was approximately 11mm per rainy day in both scenarios. Diarrhea prevalence was 5.7% (95% CI 4.4%, 7.0%) higher under SSP5, 3.4% (95% CI 2.7%, 4.3%) higher under SSP2, and 0.4% lower under SSP 1 (95% CI 0.2%, 0.9%) (Figure 4). Projected enteropathogen prevalence under SSP1 was similar with prevalence in the study period. Under SSP2, prevalence of enteric bacteria carriage was up to 33% higher, although a decrease was estimated for aEPEC. We estimated small increases in the prevalence of *E. bieneusi*, and diarrhea, and up to 13% decreases in the prevalence of *Giardia*, sapovirus and norovirus. Overall, enteropathogen carriage estimates were similar for SSP2 and SSP5. Using alternative precipitation lag periods, we generally observed the largest percent changes with 1-week lags for viruses and 2-week lags for bacteria (Figures A10-12). For parasites, the direction of estimates varied by lag period.

**Figure 4.**
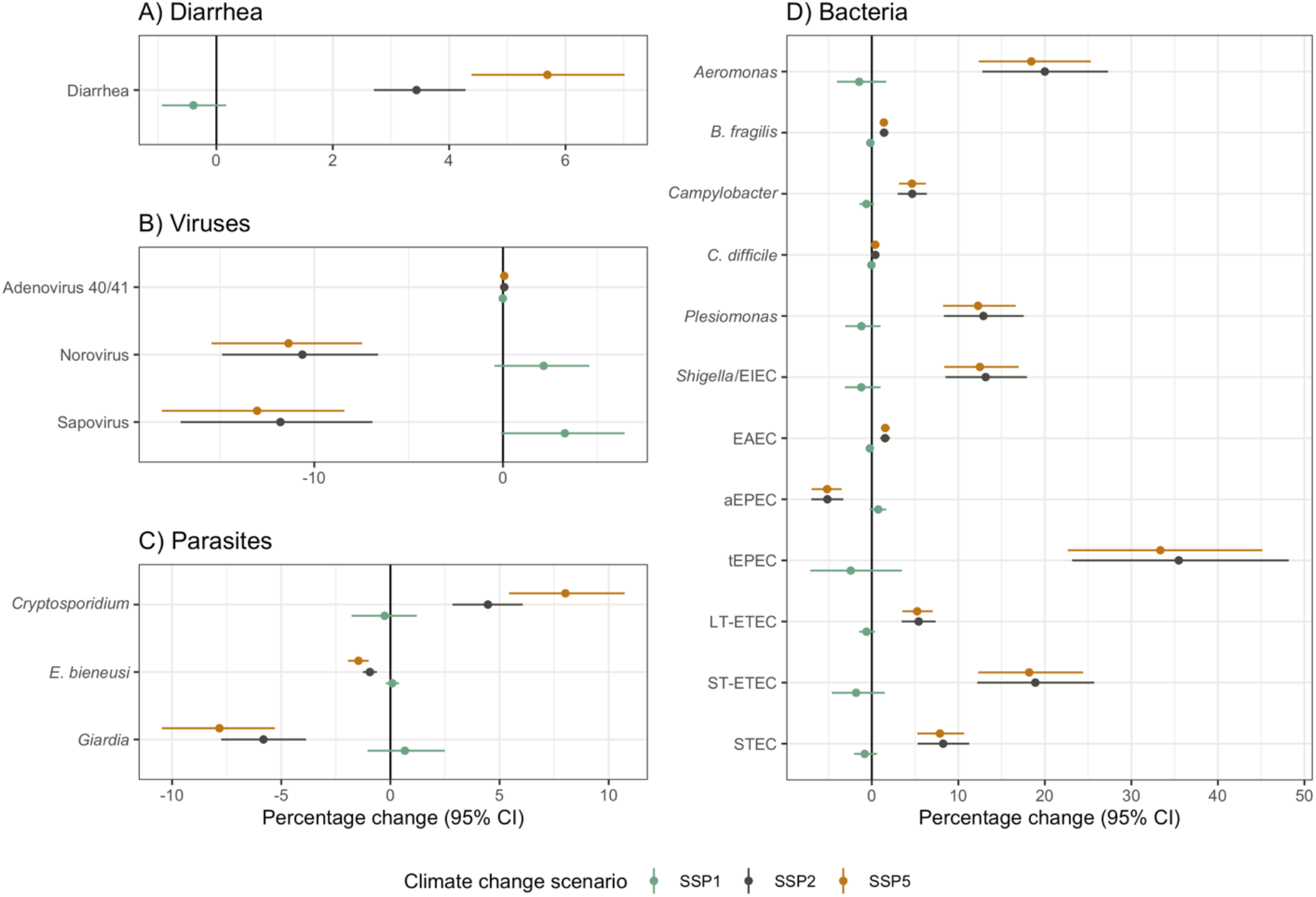
Percentage change in diarrhea and enteropathogen prevalence under climate change scenarios. Colors indicate Shared Socioeconomic Pathway scenarios: SSP1: sustainable development (SSP1); middle of the road (SSP2); and fossil fuel-based development (SSP5). Point estimates display relative percentage changes in outcome prevalence in each scenario in 2050 compared to prevalence during the study period. Error bars indicate 95% confidence intervals estimated with a clustered bootstrap with 1,000 replicates. Point estimates with small counterfactual shifts in the precipitation distribution produced narrower confidence intervals. Panel A) includes diarrhea measurements in children aged 6 months - 5.5 years in the control arms in the original trial. Panels B-D) include measurements in children approximately 14 months of age in the control, combined water + sanitation + handwashing (WASH), nutrition, and combined nutrition + WASH arms of the original trial. We present results for the lag periods that best aligned with the incubation period for each outcome (Appendix 7); results with alternative lag periods are in Figures A11-13.

### Other analyses

We did not detect effect modification in any of our pre-specified analyses. Our negative control analysis using caregiver-reported child bruising found null associations with each risk factor, suggesting that potential misclassification of the outcome did not substantially influence our results (Table A2, Figure A13).

## Discussion

We found that higher weekly average temperatures were associated with higher prevalence of diarrhea, norovirus, and STEC; above-median weekly precipitation was associated with higher prevalence of diarrhea, *Cryptosporidium*, adenovirus 40/41, and multiple enteric bacteria and lower prevalence of *Giardia*, norovirus, and sapovirus. Surface water presence was only associated with carriage of certain enteric bacteria and parasites and not with diarrhea. Under a sustainable development climate change scenario (SSP1), we projected no changes in outcome prevalence. Under middle of the road (SSP2) and fossil fuel-based development scenarios (SSP5), we projected that prevalence was up to 6% higher for diarrhea, up to 33% higher for enteric bacteria – including moderately higher *Shigella* and *Aeromonas*, which are responsible for substantial moderate-to-severe diarrhea in this setting^18^ – and up to 13% lower for sapovirus and norovirus.

### Precipitation and diarrhea

Above-median weekly precipitation with a 2-week lag was associated with up to 35% higher diarrhea prevalence, but heavy rainfall was associated with lower prevalence of diarrhea. A prior meta-analysis found both positive and negative associations between precipitation and diarrhea risk.^28^ Generally, heavy rainfall preceded by a dry period has been associated with higher diarrhea risk, while heavy rainfall preceded by a wet period has been associated with lower diarrhea risk. This may be because pathogens concentrate during dry periods and then are flushed into the environment when heavy rainfall occurs (the concentration-dilution hypothesis).^28^ We were not able to investigate associations with heavy rainfall preceded by a dry vs. wet period because almost all heavy rainfall periods were preceded by rainy days. Taken together, our findings suggest that in our study setting, even moderately higher precipitation was associated with meaningful increases in diarrhea.

### Precipitation and enteropathogens

We found that higher precipitation was associated with a higher prevalence of *Cryptosporidium*, adenovirus 40/41, *Aeromonas, Shigella*/EIEC, *Campylobacter*, EAEC, tEPEC, ST-ETEC, and STEC and lower prevalence of aEPEC, norovirus, sapovirus, and *Giardia*. A recent meta-analysis of data from 19 LMICs with tropical climates found that higher precipitation was associated with a small decrease in ETEC and *Campylobacter* prevalence and no difference in *Cryptosporidium, Shigella, Giardia*, or enteric virus prevalence.^10^ Our findings may differ due varying background levels of enteropathogen transmission and water and sanitation (WASH) infrastructure in our study compared to those in the review. Pathogen prevalence was similar for viruses, slightly higher for *Campylobacter* and *Cryptosporidium*, and lower for Giardia and *Shigella/EIEC* in our study compared to the prior meta-analysis.

### Temperature

For temperature, our estimated associations were more modest overall than those from previous studies. We found that an increase in weekly average temperature from 15 to 30° C was associated with approximately 5% higher diarrhea prevalence, 6.4% higher norovirus prevalence, and 13% higher STEC prevalence. One study estimated that a 1° C increase in mean temperature was associated with relative increases of 7% for diarrhea and bacterial diarrhea and no association with viral diarrhea.^9^ Another estimated that an increase from approximately 10-40° C in weekly average temperature was associated with higher risk of *Campylobacter*, ETEC, *Shigella, Cryptosporidium, Giardia*, and adenovirus and lower risk of sapovirus and rotavirus, and generally associations were stronger.^10^ Our more modest estimates may reflect differing background levels of enteropathogen transmission, co-varying environmental conditions, and WASH infrastructure. Our findings reinforce that relationships between temperature and enteropathogen carriage are heterogeneous within taxa. While higher water temperatures may reduce the concentration of some enteropathogens in surface water by increasing inactivation, bacteria may grow more quickly in higher temperatures,^29^ and higher temperatures could increase pathogen survival on land, in surface water, and in aquifers.^6^

### Climate

Our estimate that diarrhea prevalence would increase by 5.7% by 2050 under SSP5 (fossil-fuel based development) is similar to previous estimates of 7% in six geographic regions, 8% in Nepal, and 3-10% in the Gaza Strip;^11,13,16^ diarrhea prevalence in our study site was similar to that in the Gaza Strip and lower than that in Nepal. Under SSP2 and SSP5, we estimated 13% higher *Shigella spp*.*/*EIEC and 18-20% higher *Aeromonas* prevalence; these pathogens are responsible for 64% of moderate-to-severe diarrhea among children 12-24 months and 86% among children 24-59 months in our setting.^18^ Though we projected small decreases in norovirus and sapovirus burden under SSP2 and SSP5, in prior research, neither were strongly associated with moderate-to-severe diarrhea in children in Bangladesh.^18^ Climate change could increase or decrease individual enteropathogen prevalence due to the diversity of pathogen survival and inactivation processes across species.^6^

For all outcomes, prevalence was similar during the study period and predicted under SSP 1 (sustainable development). This is likely because the mean precipitation per rainy day was nearly the same during the study vs. under SSP1 even though there were more rainy days during the study. In settings with differing precipitation distributions under climate change, diarrhea and enteropathogen prevalence projections may be lower under SSP1.

A unique strength of our study is that we matched predictions under climate change by household location and month to capture fine-scale heterogeneity in the relationship between precipitation and each outcome. Prior studies have used single measures of association between temperature and diarrhea risk,^11,14,15,17^ but associations likely vary across settings.

Further, our study captured intra-annual heterogeneity in precipitation projections under climate change, while most previous studies used annual measures (e.g., mean annual temperature).^11,13–15,17^

### Limitations

This study was subject to several limitations. First, in 2014, the original trial did not collect data during peak rainy season (Figure 1), limiting our ability to make inferences about associations with heavy rainfall in that season. Second, we only measured enteropathogens in a subsample, so statistical power was limited for certain analyses; for this reason, we did not consider it feasible to estimate associations with taxa-specific diarrhea (e.g., bacterial diarrhea) as prior studies have done. However, measuring enteropathogen carriage (including asymptomatic and symptomatic infections) sheds light on climate-specific transmission patterns that may not be discernable when restricting to symptomatic cases. Third, the enteropathogen sample included a narrow age range (approximately 14 months); results may not generalize to other ages. Our effect modification analyses may also have had limited statistical power. Finally, relationships between climate, diarrhea, and enteropathogen infection likely vary by climate zone;^30^ our findings may not hold in settings with differing enteropathogen transmission levels, WASH infrastructure, and climatic characteristics.

There are numerous sources of uncertainty in climate change and health prediction models. We used projected precipitation from a model that best aligned with our study site, but projections elsewhere could differ substantially. Predictions are sensitive to the estimated association between climate and diarrhea,^11^ which could vary by location and over time. We were not able to predict prevalence under climate scenarios using temperature because the climate change projection temperature dataset differed substantially from temperature data during the study. Further, our 2050 predictions assume similar levels of WASH infrastructure, population density, and land use as in the study, but we expect these factors to change unpredictably over time.

## Conclusions

While diarrhea burden has decreased over time, our findings suggest that progress may be hindered by climate change in the absence of sustainable development. We observed heterogeneous potential impacts of climate change on enteropathogen carriage by pathogen taxa and species. Our findings inform prioritization of pathogen-specific interventions (e.g., Shigella vaccines) for young children in rural, South Asian settings with a similar diarrhea etiology to mitigate the impacts of climate change.

## Data Availability

All data produced in the present study are available upon reasonable request to the authors.

## Acknowledgments

Research reported in this publication was supported in part by the Bill and Melinda Gates Foundation (grant number OPPGD759 to the University of California, Berkeley and OPP1161946 to Stanford University) and the National Institute of Allergy and Infectious Diseases of the National Institutes of Health under Award Numbers K01AI141616 (Jade Benjamin-Chung) and R01AI166671 (Benjamin F. Arnold). Jade Benjamin-Chung is a Chan Zuckerberg Biohub Investigator. Anna T. Nguyen was supported by National Heart, Lung, And Blood Institute of the National Institutes of Health under award number T32HL151323. Jessica A. Grembi was supported by a Stanford University School of Medicine Dean’s Postdoctoral Fellowship. The content is solely the responsibility of the authors and does not necessarily represent the official views of the National Institutes of Health. We also acknowledge the Stanford Research Computing Center for computational resources at the Sherlock high-performance cluster.

## Role of the Funding Source

The finders of the study had no role in study design, data analysis, interpretation, writing of the manuscript, or the decision to submit the manuscript for publication.

## Authors’ Contributions

Jessica A. Grembi: conceptualization, data curation, formal analysis, investigation, methodology, project administration, software, resources, supervision, visualization, writing - original draft Anna T. Nguyen: data curation, formal analysis investigation, methodology, software, writing - review & editing Marie Riviere, Gabriella Barratt Heitmann, Arusha Patil, Tejas S. Athni: visualization, software, writing - review & editing Stephanie Djajadi: data curation, writing - review & editing Md Abdul Karim, Md Ohedul Islam, Rana Miah, Syeda L. Famida, Md Saheen Hossen, Palash Mutsuddi: investigation, data curation, resources, writing - review and editing Shahjahan Ali, Md Ziaur Rahman, Zahir Hussain, Abul K. Shoab: investigation, data curation, supervision, resources, writing - review and editing Rashidul Haque, Mahbubur Rahman, Leanne Unicomb, John M. Colford, Jr., Stephen P. Luby, Ayse Ercumen, Audrie Lin: project administration, supervision, resources, writing - review & editing Yoshika Crider, Andrew Mertens: resources, methodology, writing - review & editing Benjamin F. Arnold: methodology, software, project administration, writing - review & editing Adam Bennett: methodology, data curation, software, writing - review & editing Jade Benjamin-Chung: conceptualization, data curation, formal analysis, funding acquisition, methodology, project administration, resources, supervision, visualization, writing - original draft

## Ethics Committee Approval

The trial was approved by human subjects committees at the International Centre for Diarrheal Disease Research, Bangladesh (icddr,b; PR-11063), University of California, Berkeley (2011-09-3652), and Stanford University (25863).

## Notes

### Clinical Protocols

https://osf.io/f9cza/

### Author Declarations

The trial was approved by human subjects committees at the International Centre for Diarrheal Disease Research, Bangladesh (icddr,b; PR- 11063), University of California, Berkeley (2011-09-3652), and Stanford University (25863).

