## Appendix of Supplemental Figures and Tables for "Influence of climatic and environmental risk factors on child diarrhea and enteropathogen infection and predictions under climate change in rural Bangladesh"

### Appendix 1. Deviations from pre-analysis plan

1) We pre-specified analyses with a continuous measure of weekly average precipitation, but the distribution was extremely right-skewed. For these reasons, to capture days with heavier rainfall, we created an indicator for whether the weekly sum of precipitation was above or below the median for the study period with 1-, 2-, and 3-week lags.

   We pre-specified an analysis to investigate whether there was an interaction between precipitation and an indicator for whether the preceding 60 days had no rainfall, as has been done in prior studies. We were not able to conduct this analysis because above median and heavy rainfall periods were almost never preceded by a dry period within the prior 60 days.

2) We ran analyses for water flow accumulation, the enhanced vegetative index, land use, and population density per our pre-analysis plan. Overall, we did not observe associations with study outcomes. We excluded them from this manuscript. Results are available here: https://osf.io/yt67k/

3) In the analysis projecting prevalence under climate change scenarios, because generalized additive models generally produced linear relationships between weekly total precipitation and each outcome, to speed computation time, we used generalized linear models.

4) In the analysis projecting prevalence under climate change scenarios, it was not possible to perform an analogous analysis for temperature projections because each of the climate projection dataset's historical values for the study site during the study period differed substantially from the values we observed in the study.

#### Appendix 2. Details on study participants

The analysis was restricted to children enrolled in one of two cohorts of the WASH Benefits Bangladesh study: 1) the diarrhea cohort or 2) the enteropathogen cohort. These data span 3 years and were collected in one of six waves (enrollment, midline, and endline for main cohort; and baseline, midline and endline for the enteropathogen cohort which occurred ~6 months after each main survey visit for children enrolled in this cohort). Each collection wave lasted approximately 1 year. Approximately 54 clusters were sampled per month from the diarrhea cohort, and 14 clusters were sampled per month from the enteropathogen cohort in a spatially dependent manner as the study team moved around the study area over the course of a year.

The diarrhea cohort included index children (lives births of women pregnant at the time of enrollment), other children living within the same compound that were younger than 3 years at study enrollment, and children within the same household born after the index children. Caregiver-reported diarrhea was collected at enrollment (May 2012-July 2013), and approximately 13 and 26 months after enrollment (Sept 2013-Sept 2014 and Dec 2014-Oct 2015). Data on caregiver-reported diarrhea was collected at three additional times for the enteropathogen cohort approximately 7 (Dec 2012-Jan 2014), 18 (Nov 2013-Nov 2014), and 33 (Mar 2015-Mar 2016) months after enrollment.

The enteropathogen cohort included index children from a subsample of clusters that was evenly balanced across the control, WSH, nutrition, and N+WSH arms (allocation ratio 1:1:1:1). Clusters in each arm were selected based on logistical feasibility for specimen collection and transport to a central laboratory. Households were visited three times for biological specimen collection; the cohort consisted of 1,645 children at age 3 months, 1,978 children at age 14 months, and 2,049 children at age 28 months. Caregiver reported diarrhea and stool samples were collected from index children at all three timepoints: age 3.0 +/- 1.7 months (n = 1,090), age 14.0 +/- 2.0 months (n = 1,499), and age 28.1 +/- 1.9 months (n = 1,512). Enteropathogens were measured from stool collected when index children were approximately 14 months old.

Additional eligibility criteria are described elsewhere:

Luby SP, Rahman M, Arnold BF, et al. Effects of water quality, sanitation, handwashing, and nutritional interventions on diarrhoea and child growth in rural Bangladesh: A cluster randomised controlled trial. *The Lancet Global Health* 2018; 6: 30490–4.

Grembi JA, Lin A, Karim MA, et al. Effect of water, sanitation, handwashing and nutrition interventions on enteropathogens in children 14 months old: a cluster-randomized controlled trial in rural Bangladesh. *J Infect Dis* 2020. DOI:10.1093/infdis/jiaa549.

### Appendix 3. Detection of enteropathogens in stool

Real time PCR assays on the TaqMan Array Card and associated gene targets. Assays have been described previously and extensively validated (Liu 2013; Liu 2016). Nucleic acid was extracted with the QIAamp Fast DNA Stool mini kit (Qiagen, Hilden, Germany) with pre-treatment steps that included bead beating. AgPath One Step RT-PCR reagents were used for qPCR reactions, which were performed on ViiA 7 systems. Quantification cycles ($C_q$s) are the PCR cycle values at which fluorescence from amplification exceeds the background, which acts as an inverse metric of quantity of nucleic acid. Valid results required proper functioning of controls (the negative results of a sample are valid only when its external control MS2 is positive, $Cq < 35$; the positive results are valid only when the corresponding extraction blank is negative for the relevant targets, $Cq > 35$), and excluded data flagged by the real time PCR software, i.e., BADROX in combination with NOISE or SPIKE.

| Pathogen | Gene |
|---|---|
| **Virus** | |
| Adenovirus 40/41 | Fiber gene |
| Astrovirus | Capsid |
| Norovirus GI/GII | GI ORF1-2 and GII ORF1-2 |
| Rotavirus | *NSP3* |
| Sapovirus | *RdRp* |
| **Bacteria** | |
| Enteroaggregative *Escherichia coli* (EAEC)* | *aaiC*, *aatA* |
| Enteropathogenic *E. coli* (EPEC)* | *bfpA*, *eae* |
| Enterotoxigenic *E. coli* (ETEC)* | *LT*, *STh*, *STp* |
| Shiga toxin-producing *E. coli* (STEC)* | *stx1*, *stx2* |
| *Aeromonas* | Aerolysin |
| *Bacteroides fragilis* | EGBF |
| *Campylobacter* spp. | *cpn60* |
| *Campylobacter jejuni/coli* | *cadF* |
| *Clostridium difficile* | *tcdA*, *tcdB* |
| *Helicobacter pylori* | *ureC* |
| *Plesiomonas shigelloides* | *gyrB* |
| *Salmonella enterica* | *ttr* |
| *Shigella* spp./Enteroinvasive *E. coli* (EIEC) | *ipaH* |
| *Vibrio cholerae* | *hlyA* |
| **Fungi** | |
| *Encephalitozoon intestinalis* | SSU rRNA |
| *Enterocytozoon bieneusi* | *ITS* |
| **Protozoa** | |
| *Cryptosporidium* spp. | 18S rRNA |
| *Entamoeba histolytica* | 18S rRNA |
| *Entamoeba* spp. | 18S rRNA |
| *Giardia* spp. | 18S rRNA |
| *Cyclospora cayetanensis* | 18S rRNA |
| *Cystoisospora belli* | 18S rRNA |
| **Helminth** | |
| *Ancylostoma duodenale* | *ITS2* |
| *Ascaris lumbricoides* | *ITS1* |
| *Blastocystis* spp. | 18S rRNA |
| *Hymenolepis nana* | *ITS1* |
| *Necator americanus* | *ITS2* |
| *Strongyloides stercoralis* | Dispersed repetitive sequence |
| *Schistosoma* spp. | *ITS* |
| *Trichuris trichiura* | 18S rRNA |
| **Controls** | |
| MS2 | *MS2g1* |
| PhHV | *gB* |

* *E. coli* pathotypes were defined as follows: EAEC (*aaiC*, or *aatA*, or both), atypical EPEC (*eae* without *bfpA*, *stx1*, and *stx2*), typical EPEC (*bfpA* and *eae*), ETEC (STh, STp, or LT), STEC (*eae* without *bfpA* and with *stx1*, *stx2*, or both).

Liu J, Gratz J, Amour C, et al. A Laboratory-Developed TaqMan Array Card for Simultaneous Detection of 19 Enteropathogens. *Journal of Clinical Microbiology*. 2013;51(2):472-480. doi:10.1128/JCM.02658-12

Liu J, Gratz J, Amour C, et al. Optimization of quantitative PCR methods for enteropathogen detection. *PLoS One* 2016; 11:1–11.

#### Appendix 4. Description of risk factors and data sources

| Spatial risk factor | Source | Description | Unit of measurement | Temporal resolution | Spatial resolution |
|---|---|---|---|---|---|
| Vapor pressure deficit | Terraclimate [1] | The difference between observed water vapor pressure and the water vapor pressure at full air saturation. Low VPD is associated with humid air, while high VPD is associated with dry air. | kPa | Monthly | 4 km (1/24th degree) |
| Surface water in close proximity to household | Global Surface Water Explorer [2] | Multiple variables were used from this dataset to create the following risk factors for analysis:<br>• Proportion of area within radius (250, 500, 750m) with surface water detected:<br>○ Any surface water<br>○ Seasonal surface water<br>○ Ephemeral surface water<br>• Distance tertile from household to:<br>○ Any surface water<br>○ Seasonal surface water<br>○ Ephemeral surface water | months | Monthly | 30 meters |
| Precipitation | NOAA/OAR/ESRL Physical Science Laboratory [3,4] | The precipitation accumulation for the geographical area. Used to create variables for:<br>• Average 7-day precip, with lags<br>• Heavy rainfall in past week (total precipitation in 24h period within previous 7 days >80$^{th}$ percentile daily precipitation over the study period), with lags<br>• Rainfall categories (based on 33$^{rd}$ and 66$^{th}$ percentile cutoffs over the study period) for the 60 days preceding heavy rainfall measurement, with lags | mm; and also factor: heavy rainfall in past week (Y/N), and previous 60-day rainfall category (high, medium, low). | Daily | 55 km (0.5 degree) |
| Temperature (minimum, maximum, average) | NASA MOD11A1 [5] | A daily daytime and nighttime land surface temperature was extracted for each coordinate point throughout the study period. The absolute minimum temperature, absolute maximum temperature, and average temperature were | °C | Daily | 1 km |

| | | |
|---|---|---|
| | | reported for the 1 day, 7 days, 30 days, and 90 day periods preceding an observation. |

1 Abatzoglou JT, Dobrowski SZ, Parks SA, Hegewisch KC. TerraClimate, a high-resolution global dataset of monthly climate and climatic water balance from 1958–2015. *Sci Data* 2018; **5**: 170191.

2 Pekel J-F, Cottam A, Gorelick N, Belward AS. High-resolution mapping of global surface water and its long-term changes. *Nature* 2016; **540**: 418–22.

3 Xie P, Chen M, Yang S, *et al.* A Gauge-Based Analysis of Daily Precipitation over East Asia. *Journal of Hydrometeorology* 2007; **8**: 607–26.

4 Chen M, Shi W, Xie P, *et al.* Assessing objective techniques for gauge-based analyses of global daily precipitation. *Journal of Geophysical Research: Atmospheres* 2008; **113**. DOI:https://doi.org/10.1029/2007JD009132.

5 Wan Z, Hook S, Hulley G. MOD11A1 MODIS/Terra Land Surface Temperature/Emissivity Daily L3 Global 1km SIN Grid V006. 2015. DOI:10.5067/MODIS/MOD11A1.006.

6 M. Friedl DS-M. MCD12Q1 MODIS/Terra+Aqua Land Cover Type Yearly L3 Global 500m SIN Grid V006. 2019. DOI:10.5067/MODIS/MCD12Q1.006.

#### Appendix 5. Covariate sets for each risk factor

| Risk factor | Potential Confounder |
|---|---|
| Vapor pressure deficit | Monthly precipitation |
| Distance from surface water | Number of animals in compound |
| Frequency of surface water in close proximity to household | Monthly precipitation |
| Precipitation | None |
| Temperature | None |

In addition to the potential confounders listed above, all models were adjusted for age, sex, and household wealth quartile.

#### Appendix 6. Climate data processing

To predict future changes in diarrhea prevalence and WASH effectiveness under climate change, we pulled daily temperature and precipitation predictions for the study region in 2050 from the NEX-GDDP-CMIP6 dataset created by the NASA Center for Climate Simulations.

The dataset contains predictions of future climate conditions from 32 models, including the variations of the Norwegian Earth System Model (NorESM2), U.K. Earth System Model (UKESM), Australian Community Climate and Earth System Simulator (ACCESS), and Model for Interdisciplinary Research on Climate (MIROC).

We aimed to predict future environmental conditions using the model that best fit our observed data from the WASH Benefits trial. For each of the available models, we compared model outputs to observed temperature and precipitation distributions in the study region between 2012-2014. With the temperature models, we found that the models tracked weekly maximum temperatures well but systematically overestimated average and minimum weekly temperatures during summer months. With the precipitation models, we found that the models tracked weekly total precipitation well with the NorESM2-MM, UKESM1-0-LL, ACCESS-CM2, MIROC-ES2L among those with the best fit. Given that the climate models only fit historical data well for precipitation, we chose to exclude temperature risk factors from our analysis of expected trends under climate change.

To select the best precipitation model, we constructed variables from each set of predictions to describe total weekly precipitation under a 0-, 1-, 2- and 3- week lag from dates of diarrhea measurements. We completed this data processing to mirror the environmental risk factors we considered in assessing diarrhea prevalence and intervention effectiveness during the trials.

The mean predicted values for each model are shown in the table below. The UKESM1-0-LL model generated lagged precipitation variables that best matched the observed values in our study period between 2012-2014. We predicted values for precipitation variables in 2050 using this model for our analysis.

| **Variable** | **Trial Data** | **ACCESS-ESM1-5** | **MIROC6** | **NorESM2-LM** | **NorESM2-MM** | **UKESM1-0-LL** |
|---|---|---|---|---|---|---|
| Mean daily precipitation (mm) | 5.46 | **5.92** | 6.63 | 7.49 | 6.06 | 6.7 |
| Mean weekly total precipitation, 0 week lag | 44.13 | 41.44 | 47.6 | 52.56 | 44.79 | **44.05** |
| Mean weekly total precipitation, 1 week lag | 37.08 | 30.23 | 33.57 | 34.2 | 33.55 | **35.72** |
| Mean weekly total precipitation, 2 week lag | 35.62 | 28.02 | **35.43** | 34.59 | 32.55 | 33.92 |
| Mean weekly total precipitation, 3 week lag ( | 35.95 | 28.97 | 37.44 | 32.4 | 30.77 | **34.47** |

#### Appendix 7. Incubation periods for each pathogen outcome

The following incubation periods informed the selection of precipitation lag periods used in Figure 4.

| Pathogen | Incubation Period in Literature | Lag Used in Analysis | Citation |
|---|---|---|---|
| Adenovirus 40/41 | 3-10 days | 1 week | 11 |
| aEPEC | 9-12 hours | 1 week | 7 |
| Aeromonas | 12-48 hours | 1 week | 1 |
| B. fragilis | 1-5 days | 1 weeks | 2 |
| C. difficile | 2-3 days | 1 week | 6 |
| Campylobacter | 2-3 days | 1 week | 10 |
| Cryptosporidium | 2-10 days | 2 weeks | 4 |
| E. bieneusi | 5-12 days | 2 weeks | 9 |
| EAEC | 8-48 hours | 1 week | 7 |
| Giardia | 1-14 days | 2 weeks | 5 |
| LT-ETEC | 10-72 hours | 1 week | 7 |
| Norovirus | 1.2 days | 1 week | 8 |
| Plesiomonas | 48 hours | 1 week | 3 |
| Sapovirus | 1.7 days | 1 week | 8 |
| Shigella/EIEC | 10-18 hours | 1 week | 7 |
| ST-ETEC | 10-72 hours | 1 week | 7 |
| STEC | 1-10 days | 1 week | 7 |
| tEPEC | 9-12 hours | 1 week | 7 |

#### Figure A1. Pairwise relationships between climatic risk factors

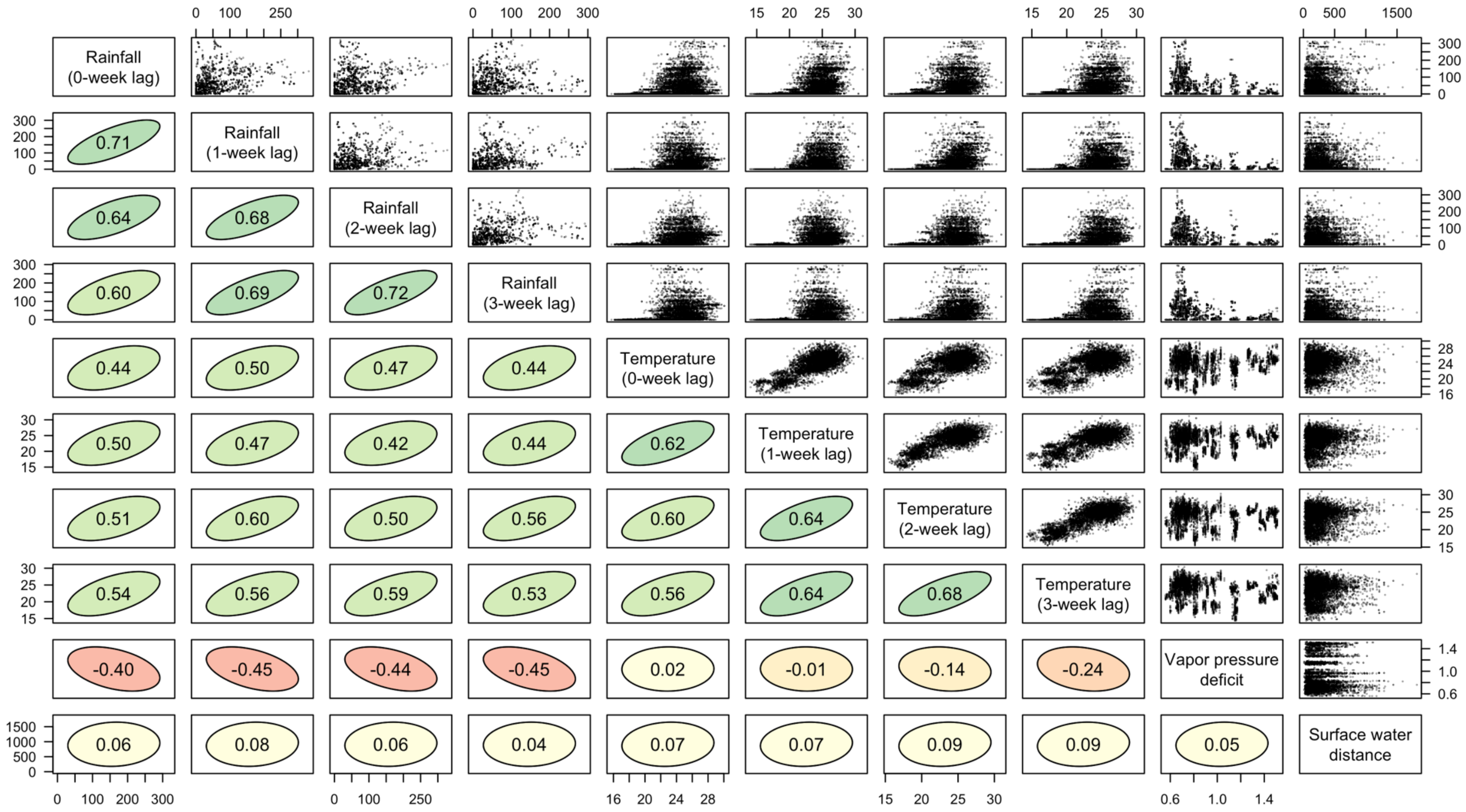

Bivariate scatter plot of continuous climatic risk factors in the diarrhea cohort. Correlation ellipses depict the strength of the association on the basis of the Spearman rank correlation, color of the ellipse indicates the direction of the correlation, and the correlation coefficient is printed inside each ellipse.

#### Figure A2. Predicted enteropathogen prevalence by weekly average temperature (C) with different lags

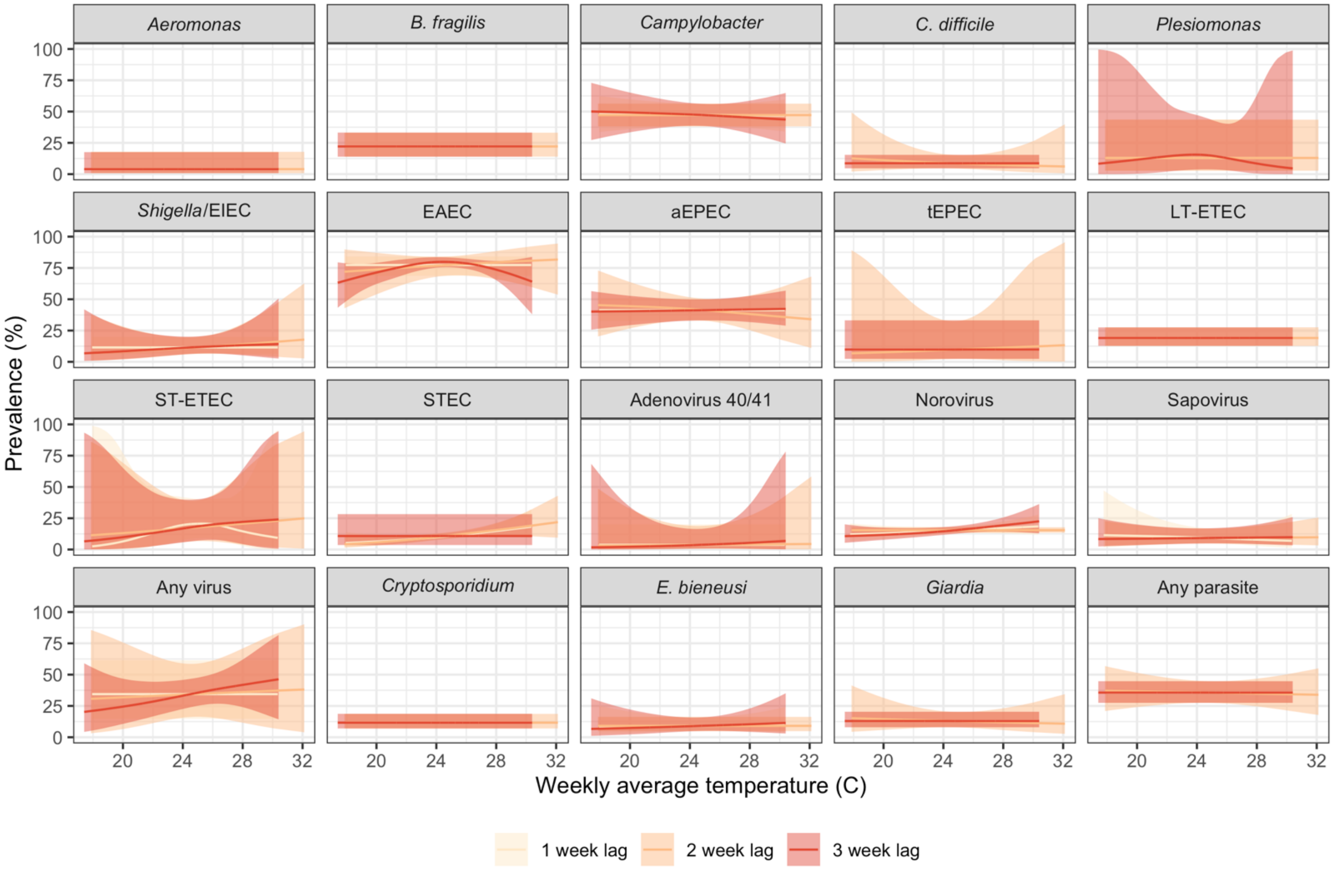

All panels present unadjusted models for children approximately 14 months of age in the control, combined water + sanitation + handwashing (WASH), nutrition, and combined nutrition + WASH arms of the original trial. Shaded bands indicate simultaneous 95% confidence intervals accounting for clustering.

#### Figure A3. Predicted diarrhea prevalence by temperature minimum, mean and maximum

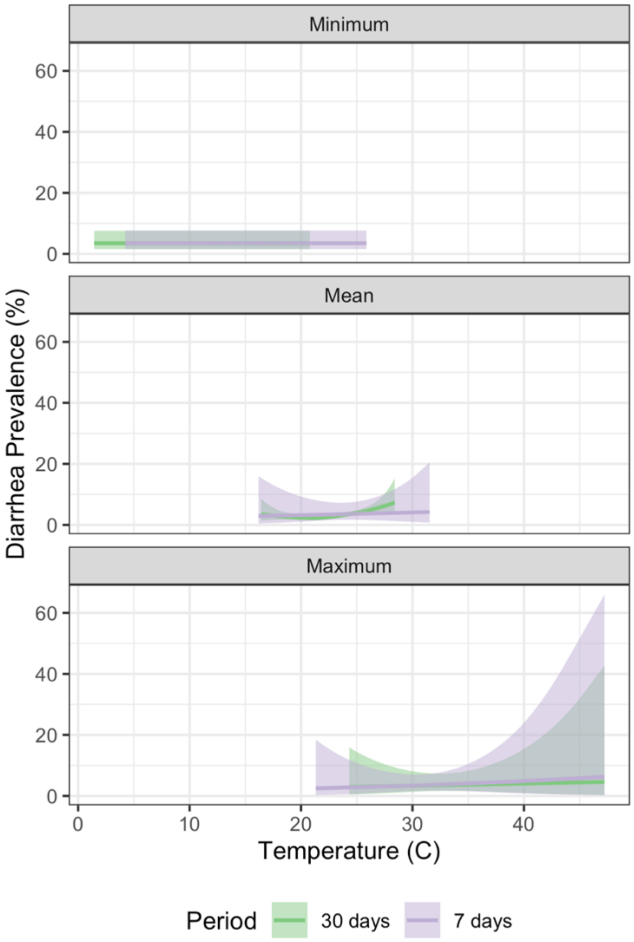

All panels present unadjusted models for diarrhea measurements in children aged 6 months - 5.5 years in the control arms in the original trial. Shaded bands indicate simultaneous 95% confidence intervals accounting for clustering.

#### Figure A4. Predicted enteropathogen prevalence by temperature minimum

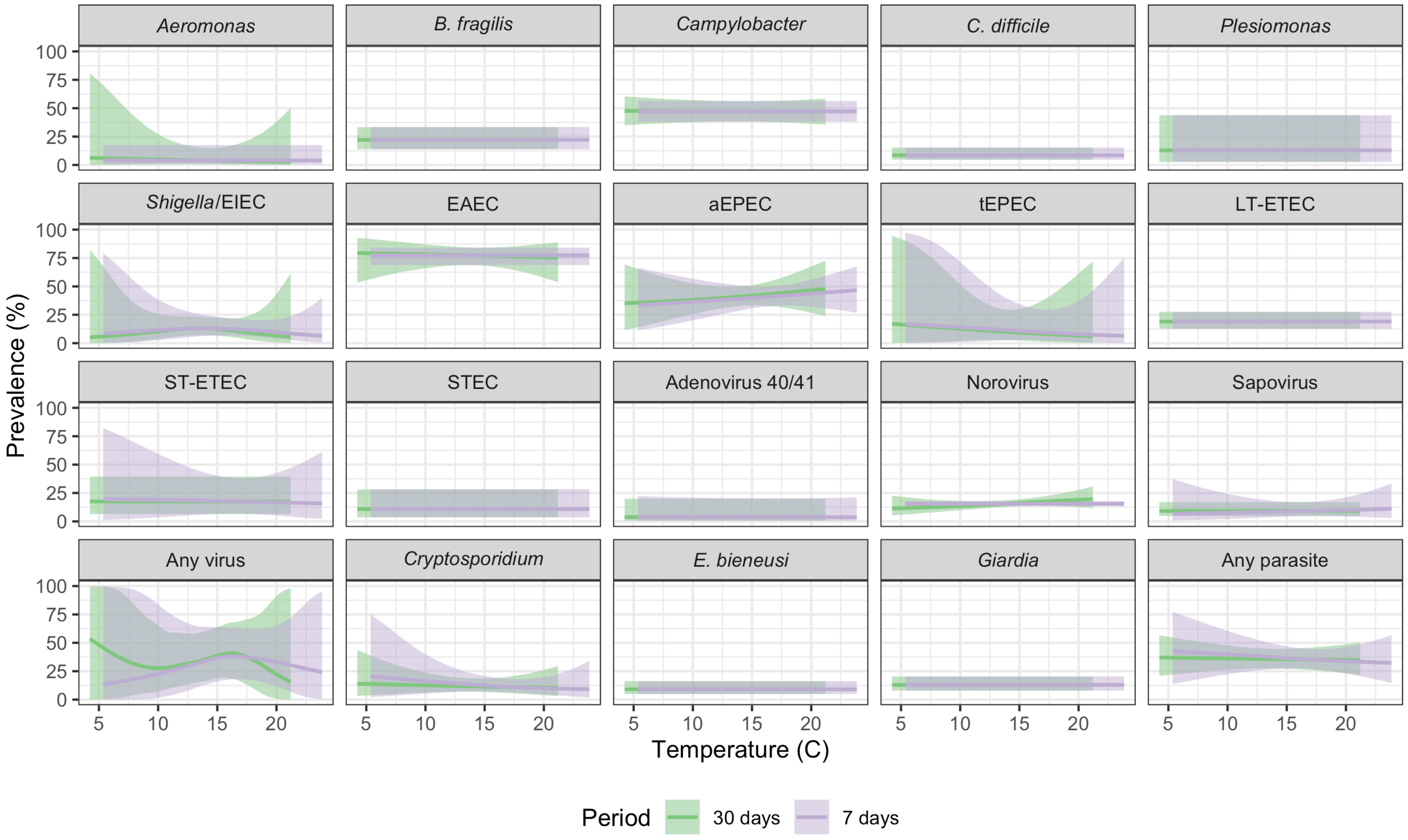

All panels present unadjusted models for children approximately 14 months of age in the control, combined water + sanitation + handwashing (WASH), nutrition, and combined nutrition + WASH arms of the original trial. Shaded bands indicate simultaneous 95% confidence intervals accounting for clustering.

#### Figure A5. Predicted enteropathogen prevalence by temperature maximum

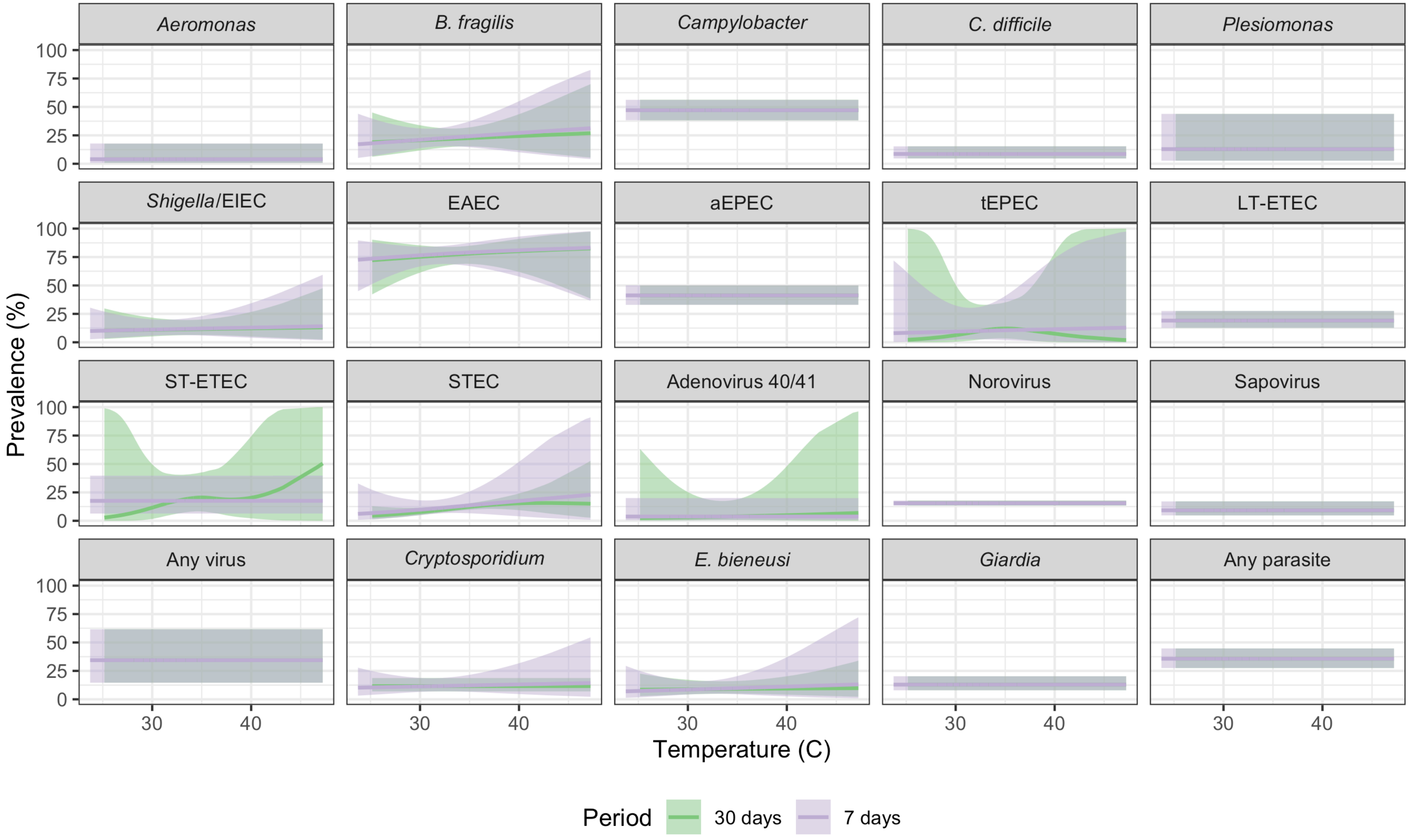

All panels present unadjusted models for children approximately 14 months of age in the control, combined water + sanitation + handwashing (WASH), nutrition, and combined nutrition + WASH arms of the original trial. Shaded bands indicate simultaneous 95% confidence intervals accounting for clustering.

### Figure A6. Diarrhea and enteropathogen prevalence by heavy rainfall

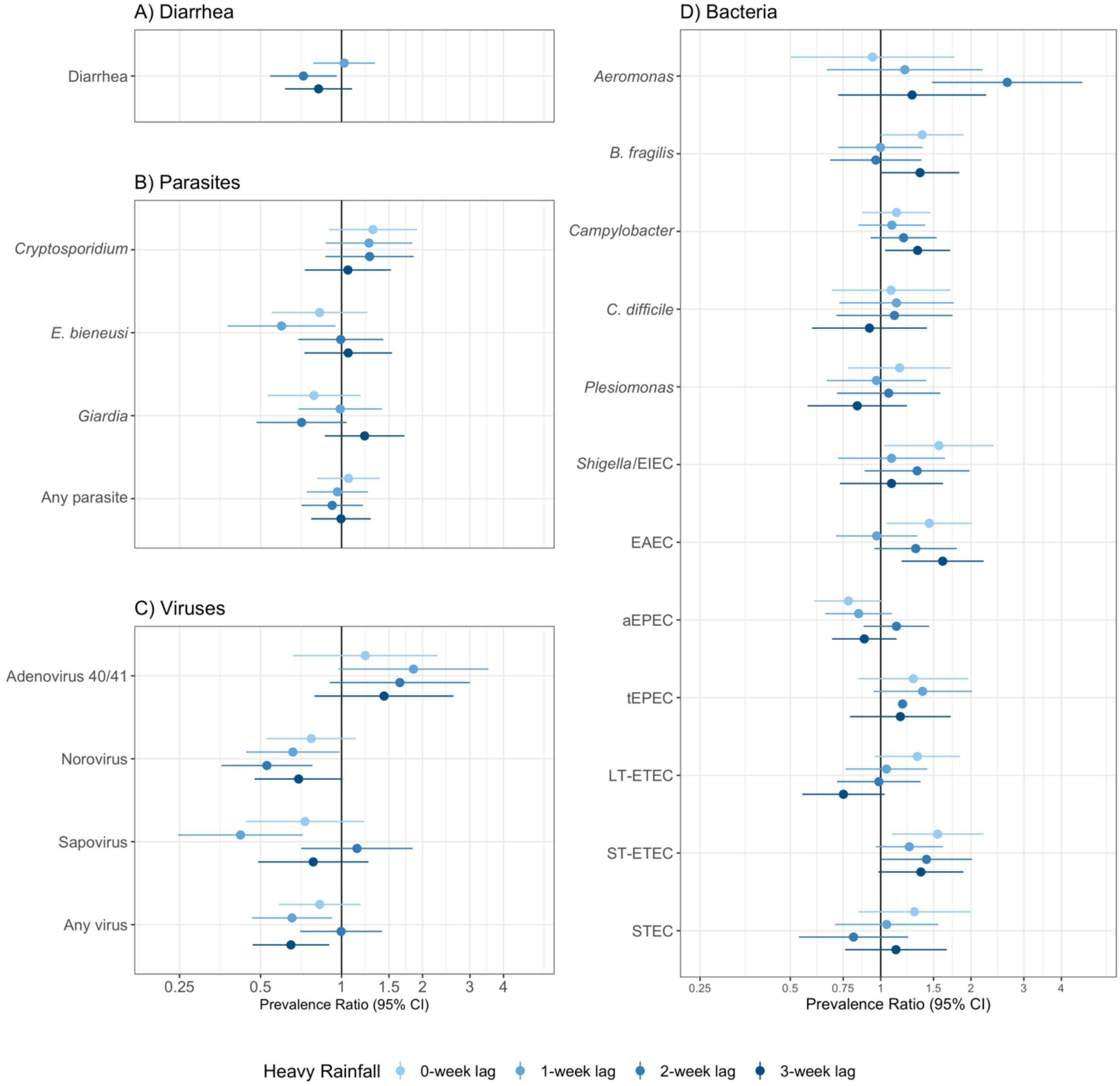

All panels present unadjusted models including an indicator variable for heavy rainfall (total weekly precipitation > 80$^{th}$ percentile during the study period); adjusted models produced similar results. Error bars present 95% confidence intervals adjusted for clustering. Panel A) includes diarrhea measurements in children aged 6 months - 5.5 years in the control arms in the original trial. Panels B-D) include measurements in children approximately 14 months of age in the control, combined water + sanitation + handwashing (WASH), nutrition, and combined nutrition + WASH arms of the original trial.

### Figure A7. Prevalence ratios for diarrhea and enteropathogen carriage and distance from study households to surface water

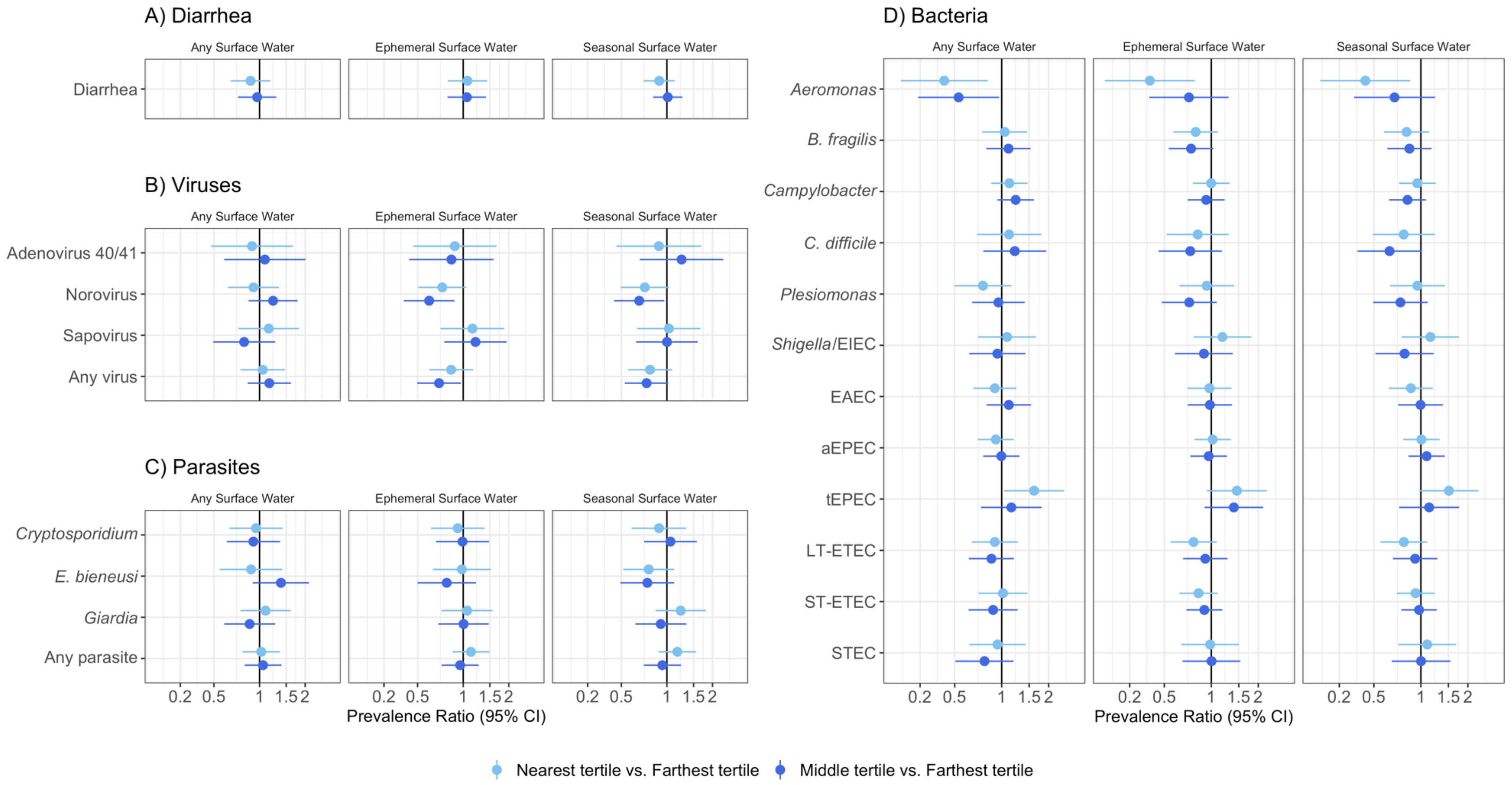

All panels present unadjusted models; adjusted models produced similar results. The independent variable was a categorical variable for tertiles of distance from each household to the nearest surface water (<165m; 165m to <316m, ≥316m). Error bars present 95% confidence intervals adjusted for clustering. Panel A) includes diarrhea measurements in children aged 6 months - 5.5 years in the control arms in the original trial. Panels B-D) include measurements in children approximately 14 months of age in the control, combined water + sanitation + handwashing (WASH), nutrition, and combined nutrition + WASH arms of the original trial.

### Figure A8. Prevalence ratios for diarrhea and enteropathogen carriage associated with the proportion of land around study households that contained surface water

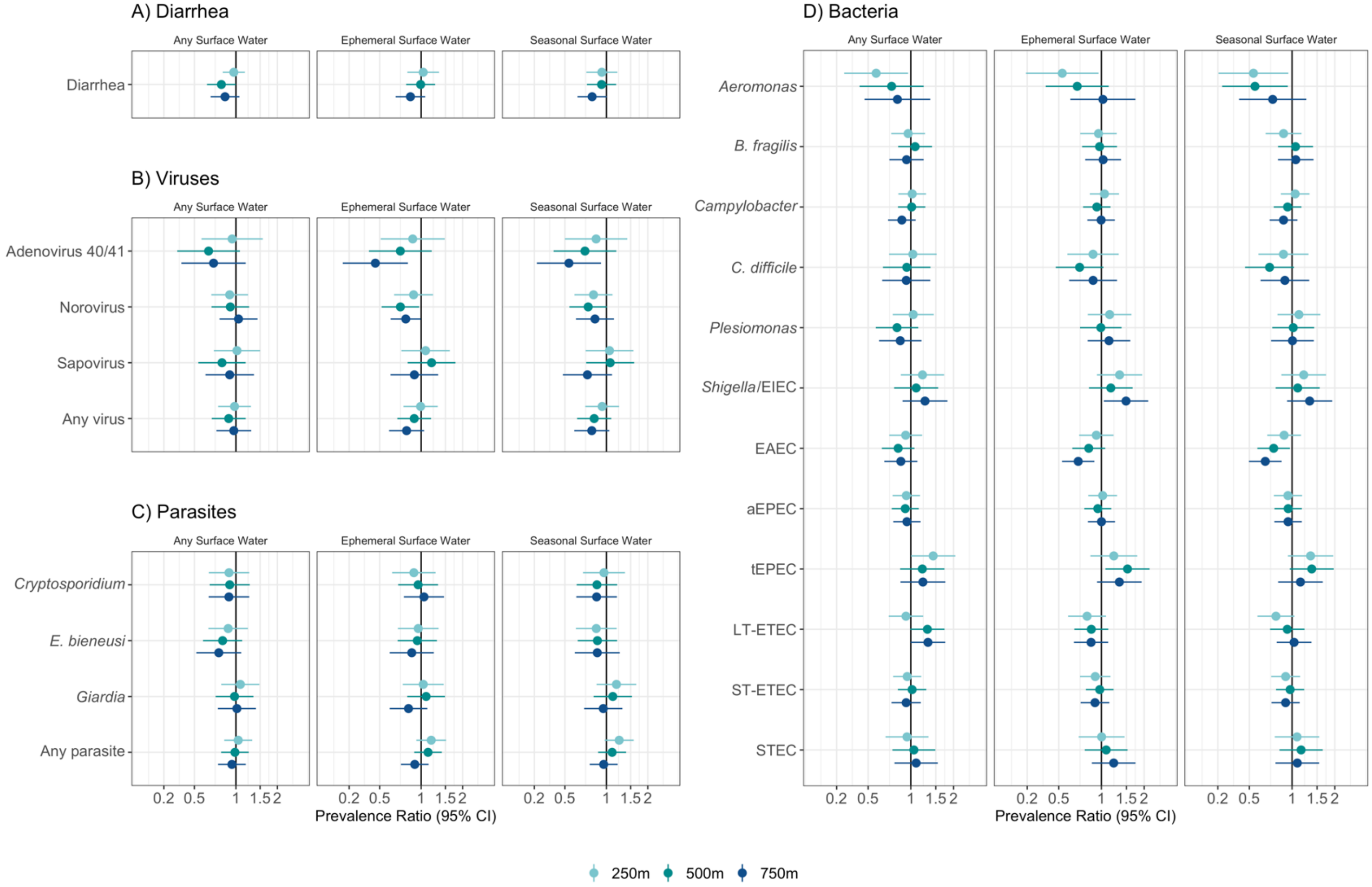

All panels present unadjusted models; adjusted models produced similar results. The independent variable was an indicator for whether the proportion of pixels with surface water within 250m, 500m, 750m of each household was above or below the median. Error bars present 95% confidence intervals adjusted for clustering. Panel A) includes diarrhea measurements in children aged 6 months - 5.5 years in the control arms in the original trial. Panels B-D) include measurements in children approximately 14 months of age in the control, combined water + sanitation + handwashing (WASH), nutrition, and combined nutrition + WASH arms of the original trial.

#### Figure A9. Predicted diarrhea and enteropathogen prevalence by vapor pressure deficit

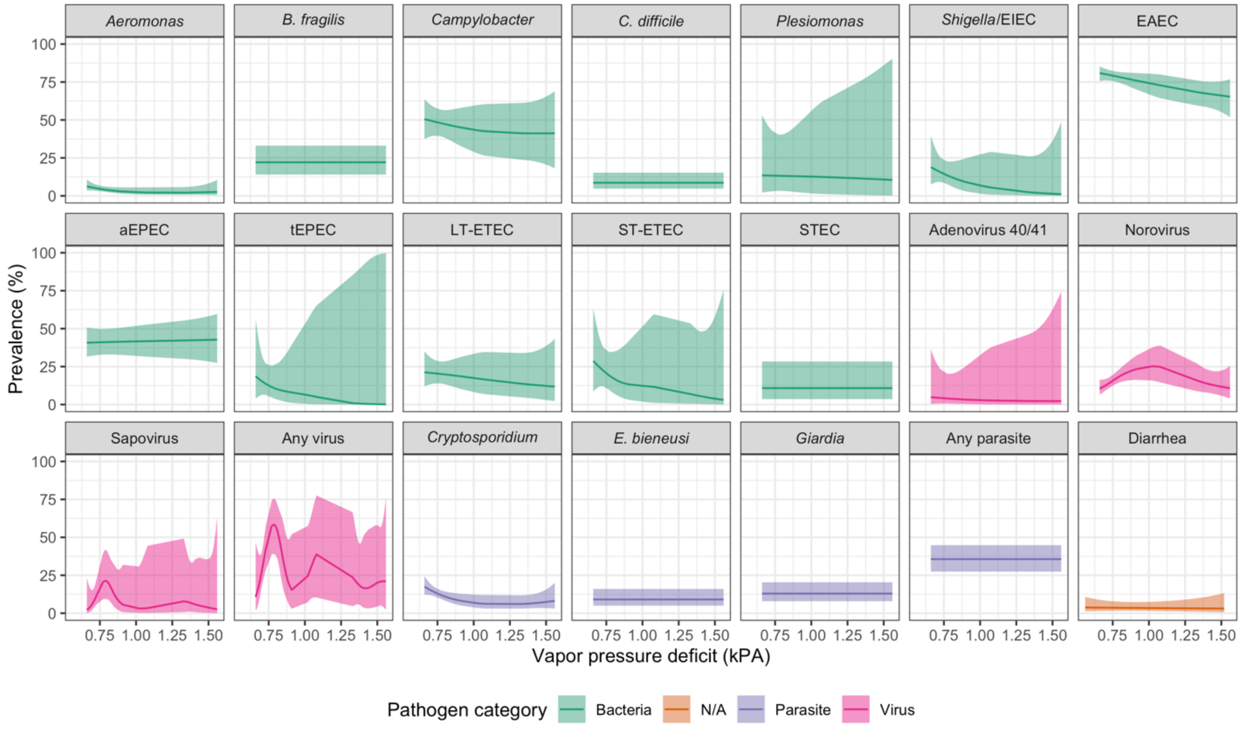

All panels present unadjusted models; shaded bands indicate simultaneous 95% confidence intervals accounting for clustering. Models included measurements in children approximately 14 months of age in the control, combined water + sanitation + handwashing (WASH), nutrition, and combined nutrition + WASH arms of the original trial.

#### Figure A10. Percentage change in diarrhea and enteropathogen prevalence under climate change scenario SSP1

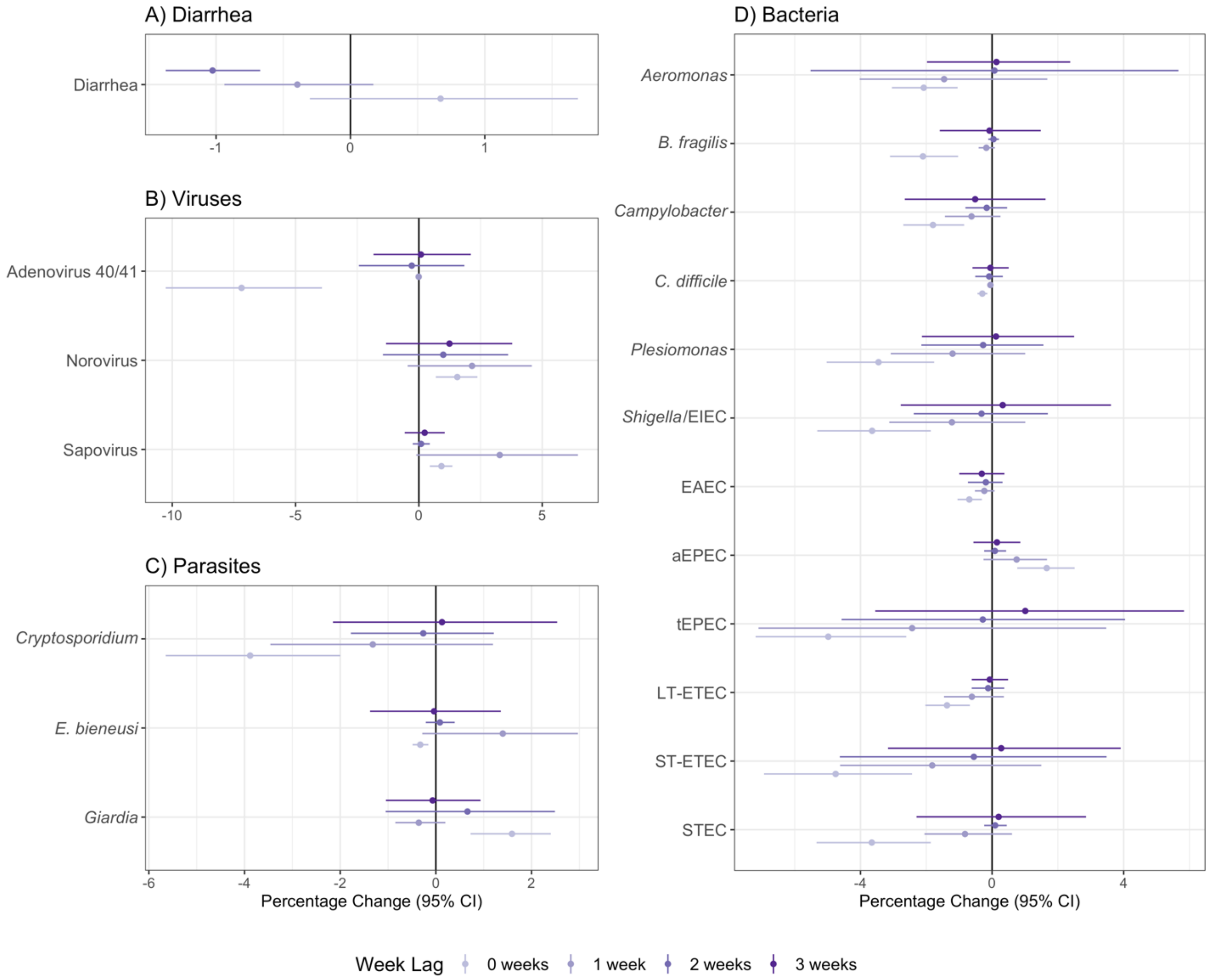

Point estimates display relative percentage changes in outcome prevalence under Shared Socioeconomic Pathway (SSP) 1 for sustainable development in 2050 compared to prevalence during the study period. Error bars indicate 95% confidence intervals estimated with a clustered bootstrap with 1,000 replicates. Point estimates with small counterfactual shifts in the precipitation distribution produced narrower confidence intervals. Panel A) includes diarrhea measurements in children aged 6 months - 5.5 years in the control arms in the original trial. Panels B-D) include measurements in children approximately 14 months of age in the control, combined water + sanitation + handwashing (WASH), nutrition, and combined nutrition + WASH arms of the original trial.

#### Figure A11. Percentage change in diarrhea and enteropathogen prevalence under climate change scenario SSP2

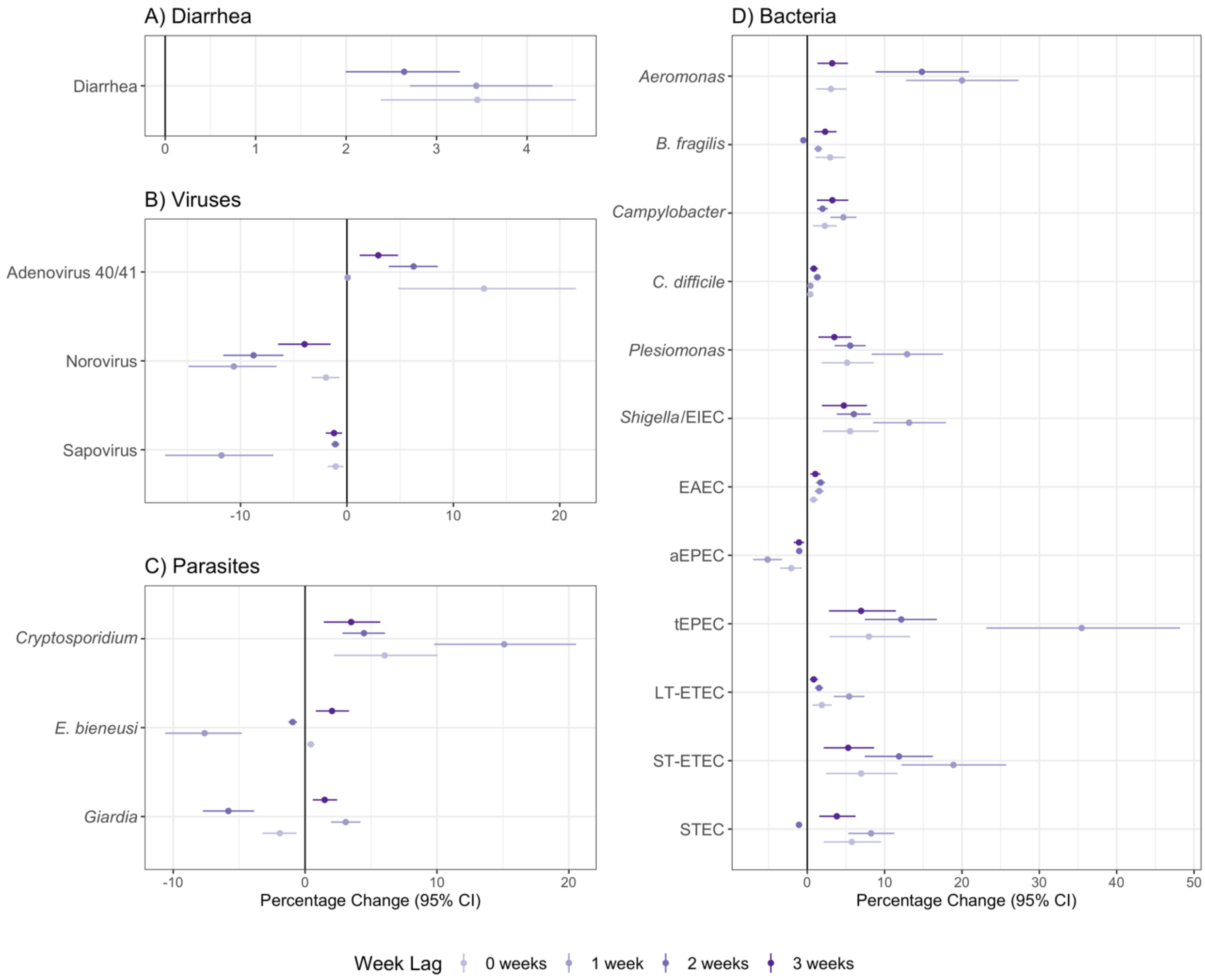

Point estimates display relative percentage changes in outcome prevalence under Shared Socioeconomic Pathway (SSP) 2 for middle of the road development in 2050 compared to prevalence during the study period. Error bars indicate 95% confidence intervals estimated with a clustered bootstrap with 1,000 replicates. Point estimates with small counterfactual shifts in the precipitation distribution produced narrower confidence intervals. Panel A) includes diarrhea measurements in children aged 6 months - 5.5 years in the control arms in the original trial. Panels B-D) include measurements in children approximately 14 months of age in the control, combined water + sanitation + handwashing (WASH), nutrition, and combined nutrition + WASH arms of the original trial.

### Figure A12. Percentage change in diarrhea and enteropathogen prevalence under climate change scenario SSP5

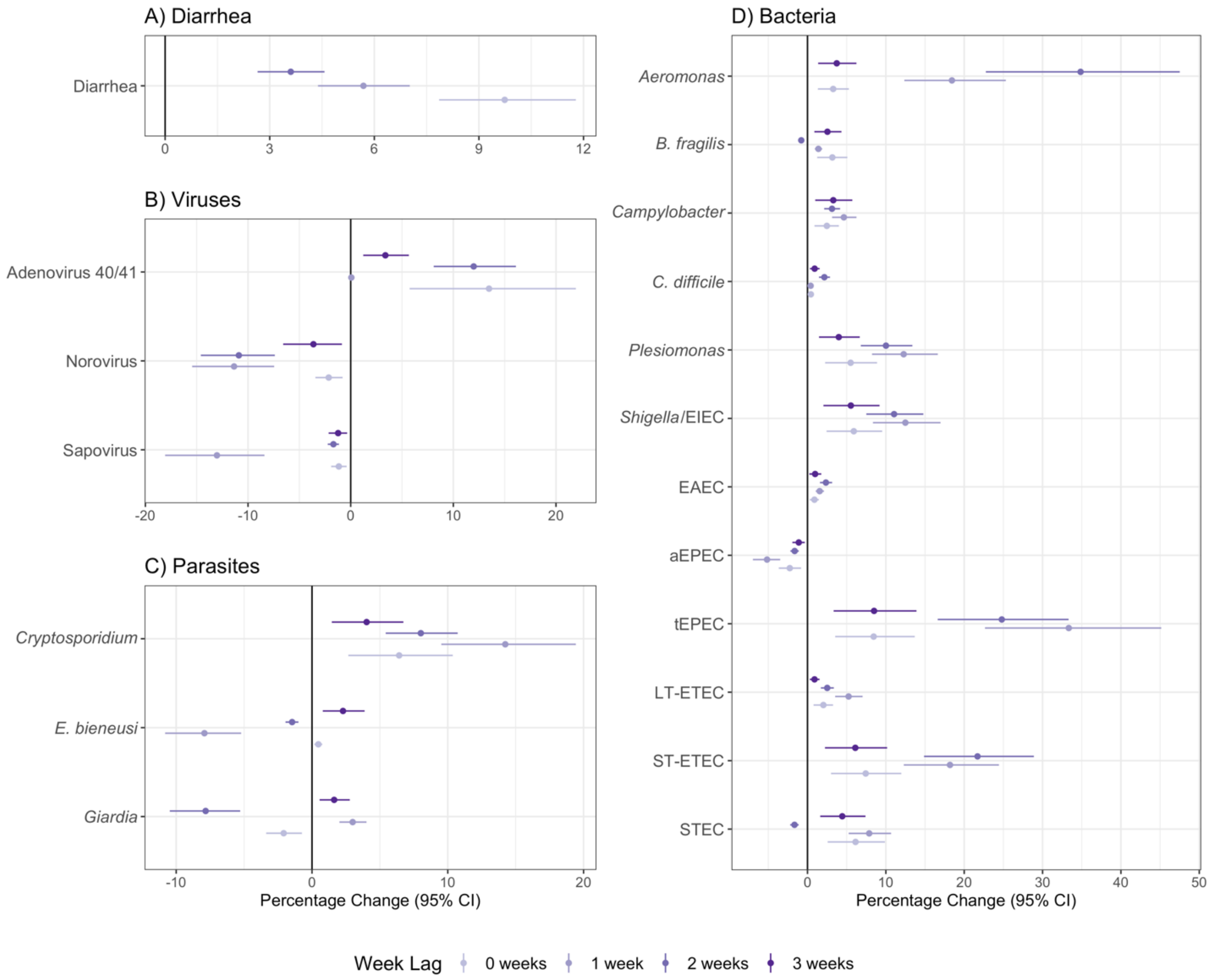

Point estimates display relative percentage changes in outcome prevalence under Shared Socioeconomic Pathway (SSP) 5 for fossil fuel-based development in 2050 compared to prevalence during the study period. Error bars indicate 95% confidence intervals estimated with a clustered bootstrap with 1,000 replicates. Point estimates with small counterfactual shifts in the precipitation distribution produced narrower confidence intervals. Panel A) includes diarrhea measurements in children aged 6 months - 5.5 years in the control arms in the original trial. Panels B-D) include measurements in children approximately 14 months of age in the control, combined water + sanitation + handwashing (WASH), nutrition, and combined nutrition + WASH arms of the original trial.

#### Figure A13. Negative control analysis – bruising and temperature

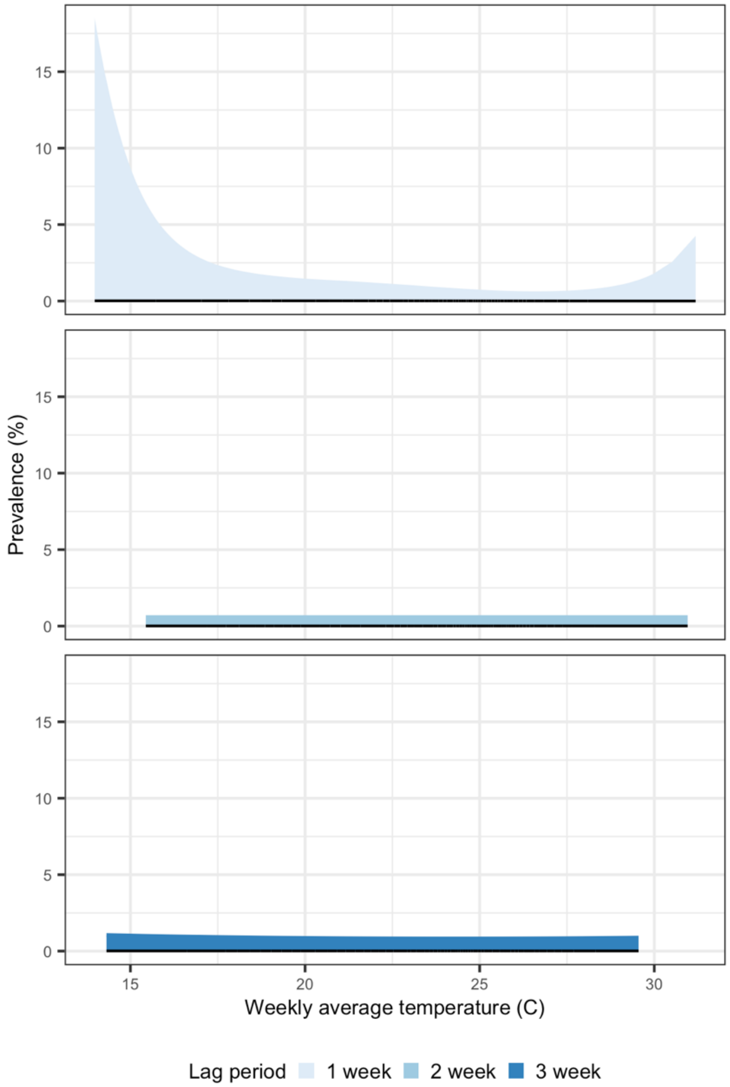

All panels present unadjusted models; shaded bands indicate simultaneous 95% confidence intervals accounting for clustering. Includes measurements in children aged 6 months - 5.5 years in the control arms in the original trial.

#### Table A1. Projected rainfall under climate change scenarios and observed rainfall during the study period

| Data Source | % of days per year with above-median weekly total rainfall | % of rainy days per year with above-median weekly total rainfall | Total annual rainfall (mm) | Total days with any rainfall |
|---|---|---|---|---|
| SSP1 - Sustainability (2050) | 37.4 | 25.8 | 1754.3 | 154.0 |
| SSP2 - Middle of the Road (2050) | 51.9 | 43.4 | 3329.1 | 186.1 |
| SSP5 - Fossil-fueled Development (2050) | 54.3 | 40.3 | 3782.1 | 190.1 |
| Trial Data Avg. (2012-2015) | 50.2 | 37.1 | 2031.5 | 185.2 |

Includes precipitation for households with children aged 6 months - 5.5 years in the control arms in the original trial.

#### Table A2. Unadjusted prevalence ratios for caregiver-reported child bruising in the prior 7 days

| Risk factor | N | Prevalence exposed | Prevalence unexposed | Unadjusted prevalence ratio (95% CI) |
|---|---|---|---|---|
| Heavy Rain | | | | |
| 1-Week Lag | 6579 | 2.58% | 3.41% | 0.81 (0.36, 1.81) |
| 2-Week Lag | 6579 | 3.37% | 3.21% | 1.05 (0.55, 1.99) |
| 3-Week Lag | 6579 | 2.33% | 3.43% | 0.68 (0.45, 1.02) |
| Above Median Weekly Sum of Precipitation | | | | |
| 1-Week Lag | 6579 | 2.97% | 3.54% | 0.76 (0.45, 1.28) |
| 2-Week Lag | 6579 | 2.92% | 3.57% | 0.82 (0.63, 1.07) |
| 3-Week Lag | 6579 | 2.94% | 3.56% | 0.87 (0.51, 1.46) |
| Distance Tertile from Any Surface Water | | | | |
| Close vs. Far | 6579 | 3.61% | 3.54% | 0.92 (0.35, 2.38) |
| Medium vs. Far | 6579 | 2.54% | 3.54% | 0.72 (0.27, 1.95) |
| Distance Tertile from Seasonal Surface Water | | | | |
| Close vs. Far | 6579 | 3.29% | 3.15% | 0.83 (0.31, 2.25) |
| Medium vs. Far | 6579 | 3.28% | 3.15% | 1.04 (0.41, 2.64) |
| Distance Tertile from Ephemeral Surface Water | | | | |
| Close vs. Far | 6579 | 3.14% | 3.20% | 0.83 (0.31, 2.19) |
| Medium vs. Far | 6579 | 3.39% | 3.20% | 1.04 (0.4, 2.67) |
| Above Median Proportion of Surface Water Near Household | | | | |
| Any Surface Water, 250m radius | 6579 | 3.22% | 3.26% | 0.89 (0.4, 1.99) |
| Seasonal Surface Water, 250m radius | 6579 | 3.40% | 3.14% | 1.08 (0.82, 1.43) |
| Ephemeral Surface Water, 250m radius | 6579 | 3.23% | 3.24% | 0.84 (0.35, 2.00) |
| Any Surface Water, 500m radius | 6579 | 3.03% | 3.46% | 0.82 (0.09, 7.56) |
| Seasonal Surface Water, 500m radius | 6579 | 3.18% | 3.30% | 0.96 (0.74, 1.26) |
| Ephemeral Surface Water, 500m radius | 6579 | 3.13% | 3.35% | 0.82 (0.46, 1.44) |
| Any Surface Water, 750m radius | 6579 | 2.95% | 3.54% | 0.83 (0.64, 1.09) |
| Seasonal Surface Water, 750m radius | 6579 | 3.16% | 3.32% | 0.95 (0.73, 1.25) |
| Ephemeral Surface Water, 750m radius | 6579 | 3.16% | 3.32% | 0.80 (0.09, 7.29) |

Includes measurements in children aged 6 months - 5.5 years in the control arms in the original trial
